# Effect of systemic glucocorticoid immunosuppression timing, dose, and duration on overall survival among immune checkpoint inhibitor recipients: a retrospective multicohort study

**DOI:** 10.1101/2025.06.23.25330022

**Authors:** Guihong Wan, Nga Nguyen, Charles Lu, Sara Khattab, Boshen Yan, Munachimso Amadife, Bonnie W. Leung, Wenxin Chen, Ahmad Rajeh, Kimberly Tang, Christopher Thang, Genevieve Boland, Kerry L. Reynolds, Kun-Hsing Yu, Alexander Gusev, Nicole R. LeBoeuf, Shawn G. Kwatra, Yevgeniy R. Semenov

## Abstract

**Background:** Systemic immunosuppression (sISP), particularly with glucocorticoids, is commonly used among patients receiving immune checkpoint inhibitor (ICI) therapy. Although recent studies have demonstrated that sISP is associated with poor outcomes in ICI recipients, there is limited information on how the timing, dose, and duration of sISP affect overall survival (OS). This multi-cohort retrospective study aims to address these gaps.

**Methods:** This study included 13,086 ICI recipients from Massachusetts General Hospital, Brigham and Women’s Hospital, and Dana-Farber Cancer Institute (MGBD) between May 31, 2015, and October 11, 2021. For independent validation, 26,172 ICI recipients were identified from the TriNetX database between April 30, 2010, and October 11, 2021, using 2:1 propensity score matching. Multivariable survival analyses were conducted with Accelerated Failure Time (AFT) models, reporting Time Ratios (TRs) and 95% Confidence Intervals (CIs), where TR<1 indicates shorter OS.

**Findings:** In the MGBD cohort, sISP within one year of ICI initiation was associated with worse OS compared to patients without sISP via landmark analysis. This association was accentuated among patients receiving sISP closer to ICI initiation, with those receiving sISP within one month of ICI initiation having the worst outcomes (TR: 0.49; 95% CI: 0.44-0.54; p<0.0001). Increased dose and duration of sISP were associated with a greater survival time loss, such that doses beyond 60 mg/day had a 40% survival time loss and durations longer than 7 days were associated with a 35% survival time loss. These findings were independently validated in the TriNetX cohort.

**Interpretation:** Three key factors are associated with significantly worse OS: sISP near ICI initiation, higher sISP dose, and longer sISP duration, regardless of indication. These findings provide clinicians with valuable information to guide sISP management among ICI recipients.

**Funding:** NIH, USA

**Panel: Research in context:** *Evidence before this study:* We reviewed PubMed and existing literature for studies on systemic immunosuppression (sISP), particularly glucocorticoids, and outcomes among immune checkpoint inhibitor (ICI) recipients. Previous research, including clinical trial studies, indicated that sISP, especially at higher doses or for managing immune-related adverse events (irAEs), was associated with poorer survival. However, studies were limited by small sample sizes, focused on single cancer types, or did not comprehensively assess the nuanced impacts of sISP timing relative to ICI initiation, dose, and duration across diverse cancer types and ICI indications. Thus, a clear understanding of these specific parameters in large, real-world cohorts was lacking.

*Added value of this study:* This multi-cohort study analyzed approximately 40,000 ICI recipients, leveraging data from a large academic medical center system (MGBD) and validating findings in a national, population-level database (TriNetX) with access to granular patient-level data. We comprehensively investigated how the timing, dose, and duration of systemic glucocorticoid immunosuppression influence overall survival across a pan-cancer population. Our large scale and detailed approach provide robust, generalizable insights into these critical factors, addressing limitations of prior, smaller, or more narrowly focused investigations.

*Implications of all the available evidence:* Our findings offer clinicians critical, actionable insights for managing systemic glucocorticoid immunosuppression in patients receiving ICI therapy. Knowing that sISP administered near ICI initiation, at higher doses, or for longer durations is associated with significantly worse overall survival can guide clinical decision-making. This allows for more informed risk-benefit discussions, strategies to minimize steroid exposure when feasible (e.g., lowest effective dose, shortest duration), and consideration of steroid-sparing alternatives, ultimately aiming to optimize survival outcomes for cancer patients on ICI therapy.

## Introduction

Immune checkpoint inhibitors (ICIs) have revolutionized cancer treatment with demonstrated overall survival (OS) benefits across multiple cancers. Currently, at least 43 indications and 20 cancer types are approved for ICI therapy.^1,2^ However, individual responses to ICIs vary, and identifying factors that influence ICI efficacy remains an active area of research. One such factor is the use of systemic immunosuppression (sISP), which may negatively impact immunotherapy outcomes due to its opposing influence on immune system signaling pathways intentionally upregulated by ICIs.

Despite this, sISP, particularly by glucocorticoids, is routine among ICI recipients experiencing inflammatory symptoms. Specifically, sISP may serve as the primary treatment for pre-existing autoimmune diseases, palliative symptom management, hypersensitivity prophylaxis, or immune-related adverse events (irAEs) among ICI recipients. The development of irAEs is common, affecting up to 70% of patients, though it is associated with improved OS in some settings.^3–6^ However, understanding sISP impact on ICI response, independent from irAE development, has been challenging due to small sample sizes^7–9^, limited follow-up^10,11^, or investigations focusing on a single cancer type.^12–14^

Recently, a meta-analysis of six clinical trials found that increasing peak corticosteroid doses for managing irAEs were associated with poorer survival among patients receiving anti-PD1 and anti-CTLA-4 therapy (HR: 1.66 for 2 mg/kg; HR: 1.21 for 1 mg/kg, comparing to 0.5 mg/kg prednisolone equivalent).^7^ Despite robust clinical trial data, these findings are often not representative of real-world clinical practice. Moreover, the generalizability of this meta-analysis is limited by its evaluation of a subset of ICI recipients who received a specific combination immunotherapy with a relatively small sample size of nearly 2,000 patients. Additionally, the study did not investigate the impact of timing or duration of corticosteroid therapy on survival. Overall, the impact of sISP dose, timing, and duration on ICI response across multiple cancers and indications remains unclear.^15^ In this study, we leverage a large multi-institutional cohort of ICI recipients to assess the impact of these factors on OS and validate our findings using an independent population-level database.

## Methods

### Study design and patients

In this retrospective study, we identified cancer patients treated with programmed cell death-1 (PD-1), programmed death ligand-1 (PD-L1), or cytotoxic T-cell lymphocyte-associated protein 4 (CTLA-4) inhibitors, either as monotherapy or combination therapy (**Table S1**). We collected two cohorts (**Figure 1**): one from Massachusetts General Hospital, Brigham and Women’s Hospital, and Dana-Farber Cancer Institute between May 31, 2015, and June 29, 2022 (**MGBD cohort**); and another from a US population-based TriNetX network between April 30, 2010, and October 11, 2021 (**TriNetX cohort**) to which MGBD does not contribute data. The Mass General Brigham Institutional Review Board approved the study (Protocol #2020P002307).

**Figure 1.**
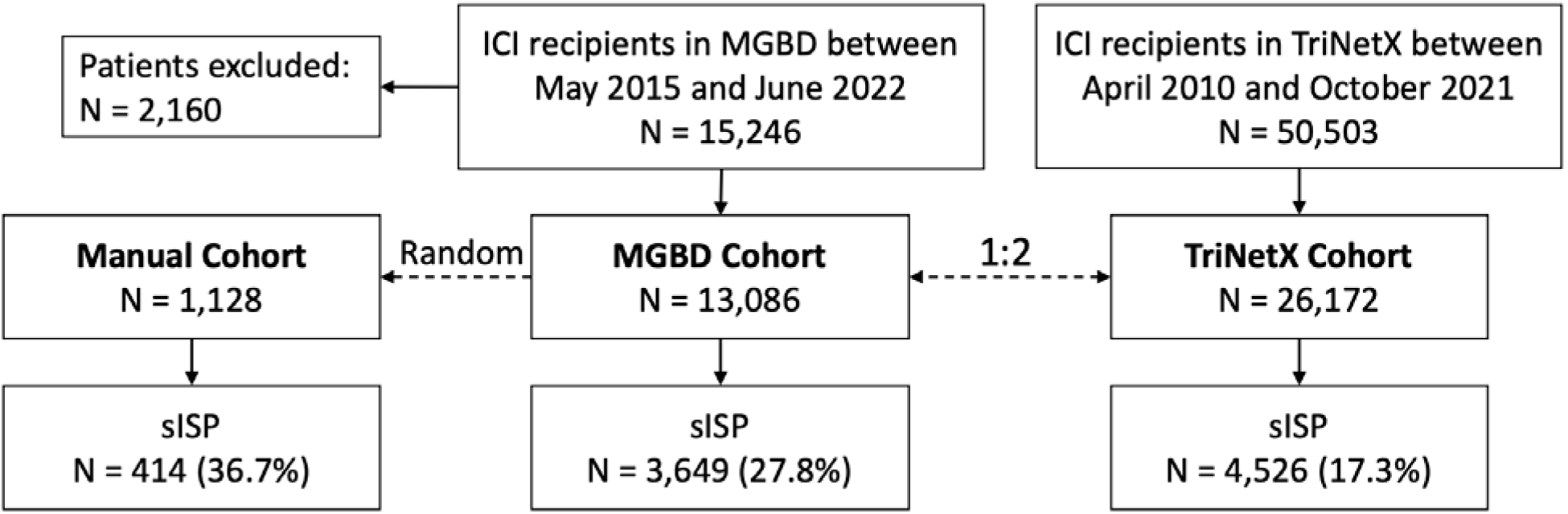
Multi-cohort study population. We identified two cohorts of individuals who received immune checkpoint inhibitor (ICI) therapy: one from Massachusetts General Hospital, Brigham and Women’s Hospital, and Dana-Farber Cancer Institute (MGBD), and another from the TriNetX database. At MGBD, a total of 2,160 patients were excluded, among which 1,482 patients with incomplete diagnosis and medication information and 678 patients receiving ICI therapy after October 11, 2021 (to be consistent with the end date with the TriNetX cohort). Following the exclusion criteria, the MGBD cohort consisted of 13,086 patients. The TriNetX cohort (N = 26,172) was identified by a 2:1 propensity score matching with the MGBD cohort by the baseline characteristics (sex, race, ethnicity, age at initiation of ICI therapy, Charlson comorbidity index, cancer type, cancer stage, ICI type, and pre-ICI treatment) as well as the year of ICI initiation. This matching aims to ensure that the two cohorts are comparable. Systemic immunosuppression (sISP) was the exposure of interest in this study. We developed a computational approach based on medication records to extract patient sISP status and validated this approach by manually phenotyping a subset of patients (N = 1,128) from the MGBD cohort. Then, we applied this validated approach to both MGBD and TriNetX cohorts. The two cohorts underwent independent analyses using the same workflow. ICI: immune checkpoint inhibitor; MGBD: Massachusetts General Hospital, Brigham and Women’s Hospital, and Dana-Farber Cancer Institute; sISP: systemic immunosuppression.

Patients with incomplete diagnosis and medication information were excluded. We also excluded patients at MGBD who received ICI therapy after October 11, 2021, to be consistent with the TriNetX cohort. The TriNetX cohort was identified by a 2:1 propensity score matching with the MGBD cohort (**Method S1**) by baseline characteristics (sex, race, ethnicity, age at initiation of ICI therapy, Charlson comorbidity index (CCI)^16^, cancer type, cancer stage, ICI type, and pre-ICI treatment) as well as year of ICI initiation (**Table S2** for definitions). The goal of this matching was not for causal effect estimation but to facilitate cohort comparability.

### Procedures

Systemic immunosuppression was the exposure of interest in this study. We developed a computational approach based on medication records to extract patient immunosuppression status (**Method S2**) and validated this approach by manually phenotyping (**Method S3**) a subset of patients (N = 1,128) from the MGBD cohort (**Manual cohort**). In our manual cohort, we observed that among the sISP cohort, all but one patient received glucocorticoid immunosuppression (**Table S3)**. Thus, for simplified and robust computational data extraction, we focused the remaining analyses on glucocorticoids with sISP henceforth referring to glucocorticoid-induced systemic immunosuppression. We applied this validated approach to both MGBD and TriNetX cohorts followed by independent analyses using the same workflow.

Potential confounding variables were identified using causal directed acyclic graph analysis (**Figure S1**) and incorporated into multivariable survival models. All variables included in this study were defined in **Table S2**. Cancer stage was estimated using secondary malignancy diagnostic codes: “distant” if secondary cancer was identified in distant sites; otherwise, “locoregional” (**Table S4**). Pre-ICI and concurrent non-ICI treatments were determined by cytotoxic chemotherapy and targeted anti-neoplastic therapy a patient received before and after ICI initiation (**Table S5**). The systemic immunosuppressive drugs included in the manual chart review are specified in **Table S6**. We used the same validated computational approach published in our recent study to identify patients who developed irAEs.^17^

### Statistical Analysis

Categorical and continuous variables were summarized using descriptive statistics. To compare two groups, we used Pearson’s chi-squared test for categorical variables and t-test for continuous variables. All statistical analyses were conducted in R version 4.1.0.

To robustly evaluate the impact of sISP timing on OS, we conducted three types of analyses for the Manual, MGBD, and TriNetX cohorts independently: **(a) Kaplan-Meier (KM) curves**: KM survival curves were utilized to examine differences among patients who received sISP within specific time windows, including [−1, 1], [−2, 2], [−3, 3], [−6, 6], [−12, 12], and [−18, 18] months, compared to the same group of patients who did not receive sISP within the [−18, 18] window (Control). The [−1, 1] group included patients who received sISP within one month before or after ICI initiation, the [−2, 2] group included patients who received sISP within two months before or after ICI initiation, and so forth. As such, the [−1, 1] group is a subset of the [−2, 2] group. **(b) Multivariable landmark analyses**: We conducted multivariable landmark analyses with Accelerated Failure Time (AFT) modeling.^18^ Landmark times were set at 1, 2, 3, 4, 5, 6, and 12 months post-ICI initiation. At each landmark time, only patients who were alive were included in the analysis. The sISP group consisted of patients who received sISP between ICI initiation and the corresponding landmark time, while the reference group included patients who didn’t receive sISP during that period. **(c) Multivariable time window analyses**: Multivariable Accelerated Failure Time (AFT) models were used to compare OS between patients who received sISP within a specific time window (e.g., [−1, 1]) versus those who did not receive sISP within this window. To account for guarantee-time bias^19^, we ensured that all patients were alive before the end date of a time window in each experiment. We also included results from Cox proportional hazards (CoxPH) models in the Appendix, although the proportional hazards assumption of CoxPH models did not hold. A sensitivity analysis focusing on sISP initiation no earlier than one month prior to ICI initiation was also performed to reflect the time period when clinicians are actively considering ICI therapy and weighing the risks and benefits of concomitant sISP therapy.

All multivariable AFT and CoxPH models were adjusted for sex, race, ethnicity, age at ICI initiation, CCI, cancer type, cancer stage, ICI type, ICI cycles, and non-ICI cancer treatment (**Table S2**). The landmark analyses were additionally adjusted for pre-ICI sISP use within three months before ICI initiation. The lognormal distribution was chosen for the AFT modeling based on Akaike’s Information Criterion (AIC) and Bayesian Information Criterion (BIC) values. For each AFT model, the Time Ratio (TR) and its 95% Confidence Interval (CI) were calculated, with TR<1 indicating a shorter survival time and TR>1 indicating a longer survival time. To enhance the robustness of our study, we conducted additional analyses for the multivariable time window analyses, where the sISP group and the no sISP group were 1:2 propensity score-matched by the same adjusted variables. Sample size calculations were conducted to verify that our analyses were appropriately powered (**Method S4**).

Sub-analyses using the Manual cohort were conducted for deeper insights. Variations in glucocorticoid dose and duration within the [−12, 12] month window were analyzed using AFT modeling, comparing each group to a common control group of patients who did not receive sISP during the period. We calculated the sISP dose as the prednisone-equivalent dose of glucocorticoids **(Table S7)**. Glucocorticoid dose was calculated as the total cumulative dose divided by the total number of days receiving sISP, stratifying patients along dose thresholds, ≥5, ≥15, ≥25, ≥35, ≥45, ≥55, ≥65, ≥75 mg/day of prednisone equivalent. The total number of days was used to determine the sISP duration, stratifying using ≥1, ≥2, ≥3, ≥5, ≥7, ≥14, ≥21, ≥28, ≥35 day thresholds. The number of patients in each sISP indication category: “Preexisting Condition and/or Other”, “Cancer and/or Other”, “irAEs and/or Other”, and “Other” was reported. Additionally, we compared patients who received sISP and those who did not among populations that developed irAEs.

## Results

### Patient Characteristics

Patient characteristics of the MGBD and TriNetX cohorts are presented in **Table S8**, including 13,086 patients at MGBD and 26,172 patients at TriNetX. Among them, 6,072 (46.4%) and 11,671 (44.6%) were females; 11,791 (90.1%) and 23,534 (89.9%) were White; 8,326 (63.6%) and 16,799 (64.2%) were less than 70 years old, respectively. The mortality rate was 7,358 (56.2%) and 10,021 (38.3%); the median duration of follow-up was 317 (IQR: 113-712) and 249 (IQR: 91-616) days, respectively. Censoring proportion in every six-month interval within the two-year follow-up duration in the two cohorts is presented in **Table S9**.

We identified 3,649 (27.8%) patients who received sISP in the MGBD cohort (**Table 1**) and 4,526 (17.3%) in the TriNetX cohort (**Table S10**). We evaluated the concordance of extracting systemic immunosuppression data between the computational method and manual chart review (**Table S11**). The overall concordance rate was 0.83 (95% CI: [0.80, 0.85]). The Positive Predictive Value (PPV) was 0.85 (95% CI: [0.81, 0.89]). The Negative Predictive Value (NPV) was 0.82 (95% CI: [0.79, 0.84]). These values were calculated for patients who underwent systemic immunosuppression within three months before or after the ICI initiation ([−3, 3]). The three-month time was selected based on its observed impact on survival outcomes.

**Table 1.** Patient characteristics of the MGBD cohort.

| Characteristics<sup>1</sup> | sISP<sup>2</sup><br>(N=3,649) | No sISP<sup>2</sup><br>(N=9,437) | P-value |
| --- | --- | --- | --- |
| **Sex** |  |  |  |
| Female | 1,739 (47.7%) | 4,333 (45.9%) | 0.076 |
| Male | 1,910 (52.3%) | 5,104 (54.1%) |  |
| **Race** |  |  |  |
| White | 3,281 (89.9%) | 8,510 (90.2%) | 0.72 |
| Asian | 123 (3.4%) | 307 (3.3%) |  |
| Black or African American | 110 (3.0%) | 254 (2.7%) |  |
| Other/Unavailable | 135 (3.7%) | 366 (3.9%) |  |
| **Ethnicity** |  |  |  |
| Not Hispanic | 3,426 (93.9%) | 8,470 (89.8%) | < 0.0001 |
| Hispanic | 100 (2.7%) | 266 (2.8%) |  |
| Unavailable | 123 (3.4%) | 701 (7.4%) |  |
| **Age at ICI Initiation (years)** |  |  |  |
| ≤60 | 1,348 (36.9%) | 2,821 (29.9%) | < 0.0001 |
| 61–70 | 1,130 (31.0%) | 3,027 (32.1%) |  |
| 71–80 | 894 (24.5%) | 2,505 (26.5%) |  |
| ≥81 | 277 (7.6%) | 1,084 (11.5%) |  |
| **Charlson Comorbidity Index** |  |  |  |
| 0 | 1 (0.0%) | 76 (0.8%) | < 0.0001 |
| 1–2 | 289 (7.9%) | 1,372 (14.5%) |  |
| 3–4 | 257 (7.0%) | 670 (7.1%) |  |
| ≥5 | 3,102 (85.0%) | 7,319 (77.6%) |  |
| **Cancer Type** |  |  |  |
| Thoracic | 875 (24.0%) | 2,450 (26.0%) | < 0.0001 |
| Male Genital/Urinary | 414 (11.3%) | 1,346 (14.3%) |  |
| Digestive | 407 (11.2%) | 1,219 (12.9%) |  |
| Melanoma | 413 (11.3%) | 905 (9.6%) |  |
| Other Skin Malignancy | 182 (5.0%) | 515 (5.5%) |  |
| Breast | 210 (5.8%) | 686 (7.3%) |  |
| Lymphoid/Hematopoietic | 305 (8.4%) | 408 (4.3%) |  |
| Female Genital | 139 (3.8%) | 497 (5.3%) |  |
| Brain/Nervous/Eye | 246 (6.7%) | 306 (3.2%) |  |
| Oral/Lip/Pharynx | 136 (3.7%) | 352 (3.7%) |  |
| Other | 322 (8.8%) | 753 (8.0%) |  |
| **Cancer Stage** |  |  |  |
| Distant | 3,082 (84.5%) | 7,170 (76.0%) | < 0.0001 |
| Locoregional | 567 (15.5%) | 2,267 (24.0%) |  |
| **Non-ICI Treatment** |  |  |  |
| Conventional Chemotherapy | 1,636 (44.8%) | 3,823 (40.5%) | < 0.0001 |
| Targeted Therapy | 513 (14.1%) | 1,258 (13.3%) |  |
| None | 1,500 (41.1%) | 4,356 (46.2%) |  |
| **ICI Type** |  |  |  |
| PD-1 | 2,678 (73.4%) | 7,227 (76.6%) | < 0.0001 |
| PD-L1 | 379 (10.4%) | 1,457 (15.4%) |  |
| CTLA-4 | 77 (2.1%) | 65 (0.7%) |  |
| Combination<sup>3</sup> | 515 (14.1%) | 688 (7.3%) |  |
| **ICI Cycles** |  |  |  |
| 1 | 788 (21.6%) | 969 (10.3%) | < 0.0001 |
| 2 | 779 (21.3%) | 1,101 (11.7%) |  |
| 3 | 607 (16.6%) | 1,277 (13.5%) |  |
| 4 | 701 (19.2%) | 2,061 (21.8%) |  |
| ≥5 | 774 (21.2%) | 4,029 (42.7%) |  |
| **Mortality Status** |  |  |  |
| Alive | 1,237 (33.9%) | 4,491 (47.6%) | < 0.0001 |
| Dead | 2,412 (66.1%) | 4,946 (52.4%) |  |
| **Duration of Follow-up (days)** |  |  |  |
| Median [IQR] | 226 [79–572] | 350 [139–751] | < 0.0001 |

<sup>1</sup> Definitions of variables are provided in Table S2.
<sup>2</sup> Systemic immunosuppression (sISP): Patients treated with systemic glucocorticoids within three months before or after initiation of ICI therapy ([-3, 3]) were considered in the sISP group; otherwise, in the no sISP group.
<sup>3</sup> Combination therapy of CTLA-4 and PD-1/PD-L1.
ICI: immune checkpoint inhibitor; sISP: systemic immunosuppression; PD-1: programmed death receptor-1; PD-L1: programmed death-ligand 1; CTLA-4: cytotoxic T-lymphocyte-associated protein 4; IQR: interquartile range.

Within the MGBD cohort (**Table 1**), the mortality rate was 2,412 (66.1%) versus 4,946 (52.4%) (p<0.0001), and the median duration of follow-up was 226 (IQR: 79-572) versus 350 (IQR: 139-751) days (p<0.0001) in the sISP and no sISP groups, respectively. Within the TriNetX cohort (**Table S10**), the mortality rate was 1,970 (43.5%) versus 8,051 (37.2%) (p<0.0001), and the median duration of follow-up was 125 (IQR: 50-336) versus 285 (IQR: 107-663) days (p<0.0001) in the sISP and no sISP groups, respectively.

### Timing of System Immunosuppression

**Figure 2** presents the KM curves within three years of follow-up after ICI initiation. For the MGBD cohort (**Figure 2A**), the KM curves demonstrated that the sISP groups consistently had poorer OS than the control group of patients who did not receive sISP within 18 months before or after ICI initiation. The steepest drop-off of OS probability was observed among patients who received sISP within one month before or after ICI initiation. The TriNetX cohort (**Figure 2B**) achieved generally consistent results. **Figure S2** presents KM curves with extended follow-up for the Manual, MGBD, and TriNetX cohorts, all of which demonstrate consistent results.

**Figure 2.**
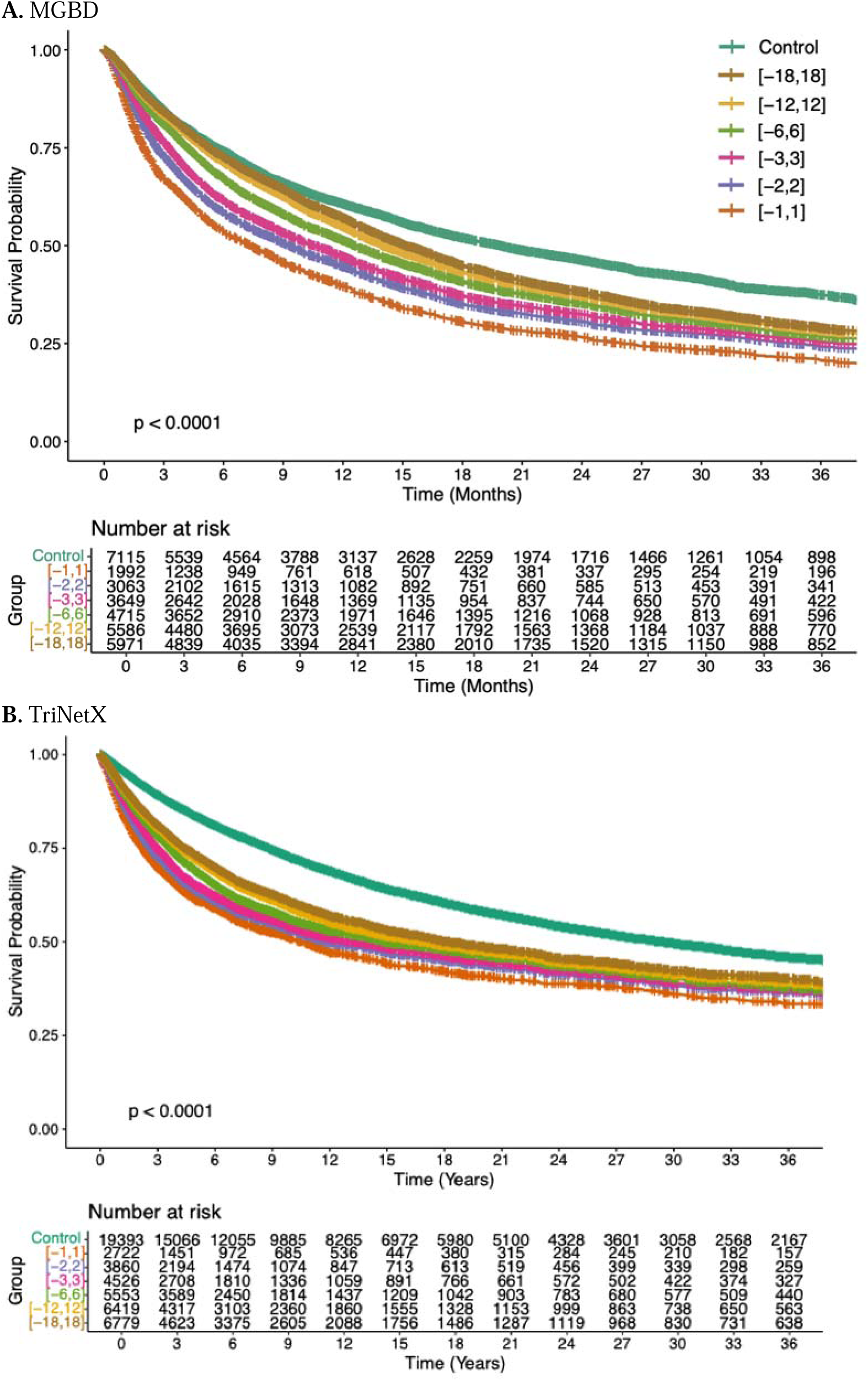
Kaplan-Meier curves by time windows of systemic glucocorticoid immunosuppression with three years follow-up. Overall, there was a decrease in survival probability as systemic glucocorticoid immunosuppression administered closer to the initiation of ICI therapy for both cohorts (p<0.0001). The “Control” group included patients who did not receive systemic glucocorticoid immunosuppression within ±18 months of the initiation of ICI therapy. The study groups were categorized by systemic glucocorticoid immunosuppression timing relative to the initiation of ICI therapy, where intervals like [−12,12] denote patients who received systemic glucocorticoid immunosuppression within ±12 months of the initiation of ICI therapy, and patients in a bigger time window (e.g., [−12,12]) is a superset of patients in a smaller window (e.g., [−6,6]). ICI: immune checkpoint inhibitor; MGBD: Massachusetts General Hospital, Brigham and Women’s Hospital, and Dana-Farber Cancer Institute.

**Figure 3** (MGBD and TriNetX) and **Figure S3** (Manual) present the results of the landmark survival analyses. The trends across three cohorts were consistent and demonstrated that patients who received sISP had poor OS, with the most pronounced negative effect observed in patients who received sISP within two months before or after ICI initiation (p <0.0001). For the MGBD cohort, the TRs at the one-, two-, and three-month landmark times were 0.56 (95% CI 0.52-0.62), 0.74 (95% CI 0.68-0.79), and 0.85 (95% CI 0.80-0.90), respectively. For the TriNetX cohort, the TRs at the one-, two-, and three-month landmark times were 0.47 (95% CI 0.43-0.52), 0.58 (95% CI 0.53-0.63), and 0.66 (95% CI 0.61-0.71). For the Manual cohort, the TRs at the one-, two-, and three-month landmark times were 0.53 (95% CI 0.37-0.78), 0.61 (95% CI 0.45-0.82), and 0.71 (95% CI 0.54-0.92).

**Figure 3.**
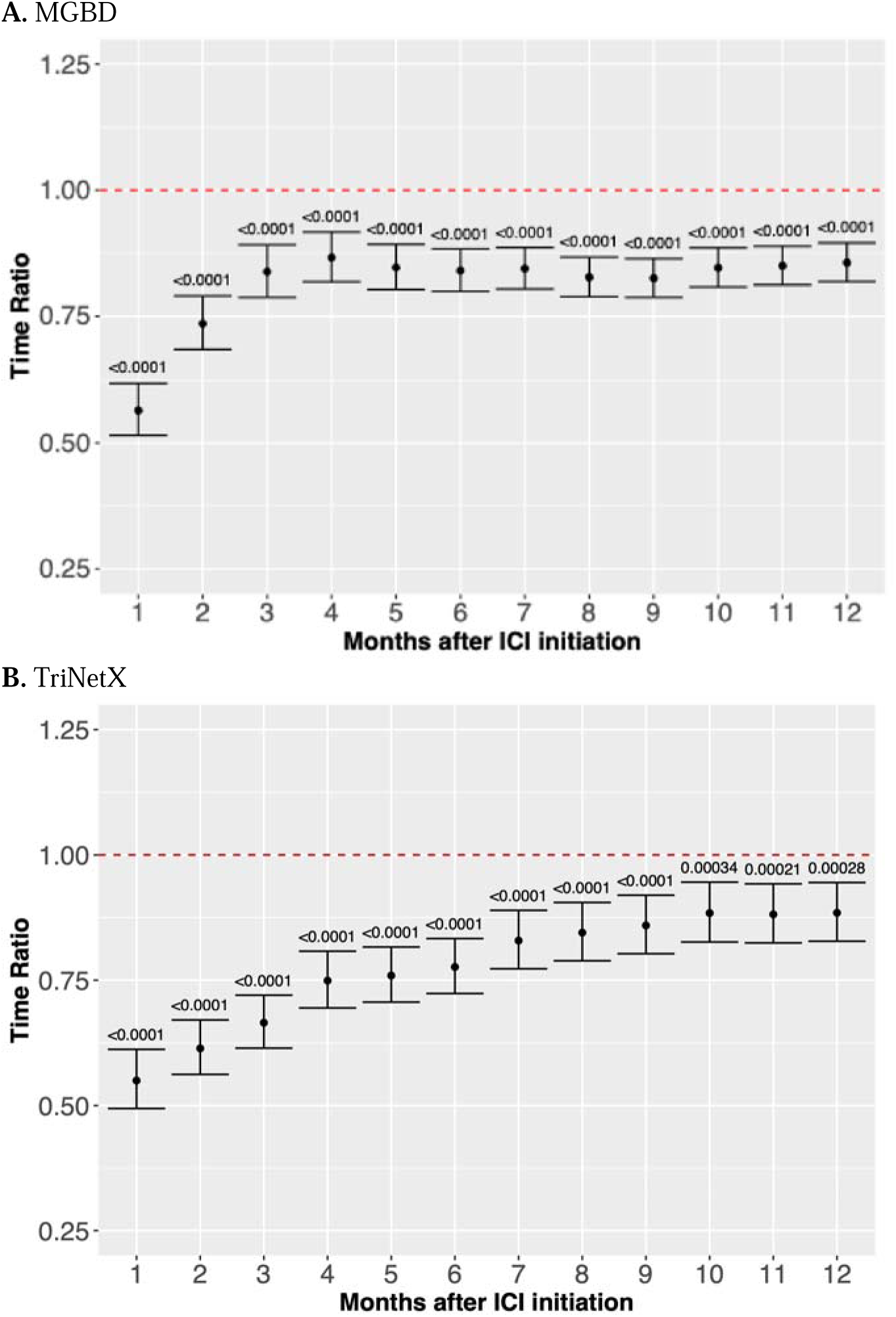
Landmark survival analysis for the associations of systemic glucocorticoid immunosuppression with overall survival. The results include Time Ratios (TRs), 95% Confidence Intervals, and significance levels, measured at various landmark times following the initiation of immune checkpoint inhibitor (ICI) therapy. Separate multivariable Accelerated Failure Time (AFT) models, adjusted for sex, race, ethnicity, age at ICI, Charlson comorbidity index, cancer type, cancer stage, non-ICI treatment, ICI type, and ICI cycles, were used at different landmark times. In each analysis, the reference group corresponded to patients who did not receive systemic immunosuppression within the landmark time. **A** and **B** show results for the MGBD and TriNetX cohorts, respectively, demonstrating that systemic immunosuppression is associated with shorter survival (TR<1 and p<0.0001). TR: Time Ratio; 95% CI: 95% Confidence Interval; ICI: immune checkpoint inhibitor; AFT: Accelerated Failure Time.

**Table 2** presents the results of the multivariable time window analyses, showing poorer OS when sISP was administered closer to ICI initiation. In both cohorts, sISP was associated with reduced survival time in every time window. In the MGBD cohort, the worst OS was observed when sISP was given within one month before or after ICI initiation (TR: 0.49, 95% CI: 0.44-0.54, p<0.0001). At increasing time windows in months, sISP administration was consistently associated with poorer OS (TR: 0.67, 95% CI: 0.62-0.74, p<0.0001 for [−2, 2]; TR: 0.75, 95% CI: 0.69-0.83, p<0.0001 for [−3, 3]; TR: 0.78, 95% CI: 0.72-0.85, p<0.0001 for [−4, 4]; TR: 0.76, 95% CI: 0.70-0.83, p<0.0001 for [−5,5]; TR: 0.75, 95% CI: 0.69-0.82, p<0.0001 for [−6, 6]), TR: 0.71, 95% CI: 0.64-0.79, p<0.0001 for [−12, 12]). In the TriNetX cohort, a similar trend was observed (TR: 0.44, 95% CI: 0.40-0.49, p<0.0001 for [−1,1]; TR: 0.55, 95% CI: 0.50-0.60, p<0.0001 for [−2,2]; TR: 0.62, 95% CI: 0.56-0.68, p<0.0001 for [−3, 3]; TR: 0.70, 95% CI: 0.64-0.78, p<0.0001 for [−4,4]; TR: 0.74, 95% CI: 0.66-0.81, p<0.0001 for [−5,5]; TR: 0.75, 95% CI: 0.67-0.83, p<0.0001 for [−6,6], TR: 0.86, 95% CI: 0.75-0.99, p=0.041 for [−12, 12]). As mentioned above, we chose AFT to model survival outcomes as some of the assumptions of Cox PH modeling were not satisfied in this data, despite demonstrating overall consistent findings with the AFT results (**Table S12)**. The results restricted to patients who received sISP no earlier than one month prior to ICI initiation are presented in Table S13, demonstrating an overall similar trend.

**Table 2:** Effect of systemic glucocorticoid immunosuppression on overall survival using multivariable AFT modeling.

|  |  | Time Window from ICI Initiation (months) |  |  |  |  |  |  |
| --- | --- | --- | --- | --- | --- | --- | --- | --- |
| Cohort |  | [-12, 12] | [-6, 6] | [-5, 5] | [-4, 4] | [-3, 3] | [-2, 2] | [-1, 1] |
| MGBD | TR | 0.71 | 0.75 | 0.76 | 0.78 | 0.75 | 0.67 | 0.49 |
|  | 95% CI | [0.64, 0.79] | [0.69, 0.82] | [0.70, 0.83] | [0.72, 0.85] | [0.69, 0.83] | [0.62, 0.74] | [0.45, 0.54] |
|  | P-value | < 0.0001 | < 0.0001 | < 0.0001 | < 0.0001 | < 0.0001 | < 0.0001 | < 0.0001 |
|  | Sample size | 5,963 | 8,586 | 9,107 | 9,689 | 10,356 | 11,190 | 12,155 |
|  | Required Sample size | 1,100 | 1,600 | 1,880 | 2,100 | 1,800 | 980 | 300 |
| TriNetX | TR | 0.86 | 0.75 | 0.74 | 0.70 | 0.62 | 0.55 | 0.44 |
|  | 95% CI | [0.75, 0.99] | [0.67, 0.83] | [0.66, 0.81] | [0.64, 0.78] | [0.56, 0.68] | [0.50, 0.60] | [0.40, 0.49] |
|  | P-value | 0.041 | < 0.0001 | < 0.0001 | < 0.0001 | < 0.0001 | < 0.0001 | < 0.0001 |
|  | Sample size | 10,330 | 15,398 | 16,613 | 18,032 | 19,633 | 21,468 | 23,562 |
|  | Required Sample size | 7,180 | 2,820 | 1,860 | 1,800 | 800 | 620 | 400 |

Adjusted Time Ratios (TRs): TR < 1 means that the covariate (e.g., systemic glucocorticoid immunosuppression) is associated with shorter survival; TR > 1 means that the covariate is associated with longer survival.
Adjusted TRs were derived from multivariable log-normal AFT models and represented their exponentiated coefficients. The AFT models were adjusted by sex, age at ICI initiation, race, ethnicity, cancer type, cancer stage, Charlson comorbidity index, ICI type, and ICI cycles.
AFT: Accelerated Failure Time; TR: Time Ratio; CI: Confidence Interval; ICI: immune checkpoint inhibitor.

The minimum sample size required for each analysis and the reasonably narrow confidence intervals (**Table 2**) indicated that our models were adequately powered. **Tables S14-S17** provide details of the models for the [−1, 1] and [−3, 3] windows for the two cohorts. **Tables S18-S19** present the results for the Manual cohort and the additional matched analyses, reaching consistent results.

**Table S20** presents the percentages of patients in the Manual cohort, categorized by indication for sISP administration across the time windows from [−1,1] to [−6,6]. The majority of patients (>50%) were receiving sISP for cancer associated symptom palliation or irAE treatment.

### Dose and Duration of Immunosuppression

Our results demonstrate a dose-dependent relationship between immunosuppression use and OS in the Manual cohort (**Figure 4A**). There is an approximately 20% reduction in OS when using immunosuppressants of ≥5 mg daily, gradually approaching an approximately 40% reduction when using ≥75 mg daily, compared to patients who did not receive sISP within the [−12, 12] month window. **Figures S4A** and **S5A** present the percentage of each immunosuppression indication, stratified by the dose category.

**Figure 4.**
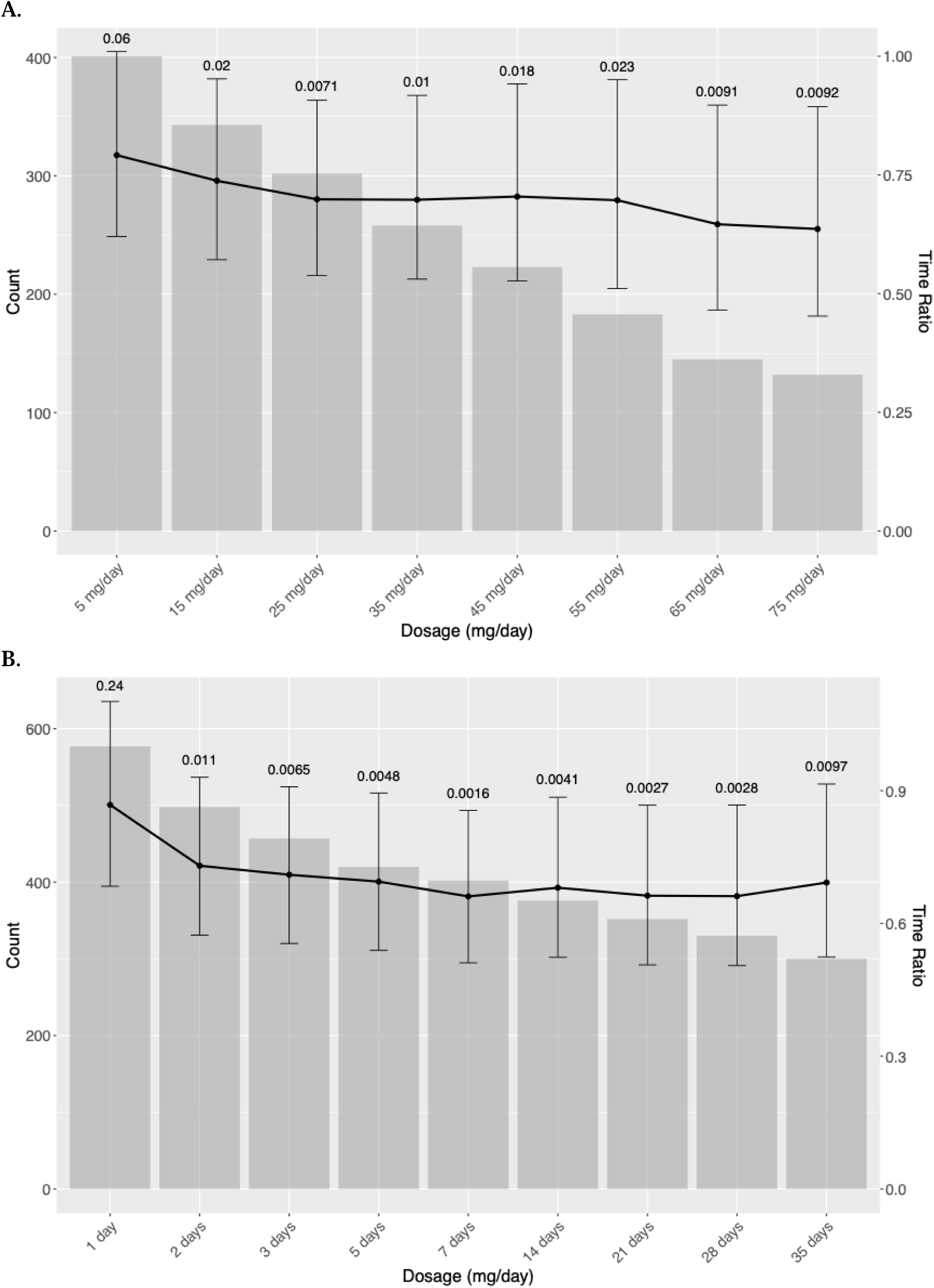
Effect of dosage and duration of glucocorticoid immunosuppression on overall survival using the manual cohort. **A.** Glucocorticoid dosage on overall survival. The x-axis represents glucocorticoid immunosuppression dosage categories in prednisone equivalent (mg/day). Each gray bar indicates the count of patients in each dosage category, and the black line represents the time ratio. The results indicate a trend of decreasing survival as dosage increases, with smaller time ratios observed in higher dosage groups. **B.** Glucocorticoid duration on overall survival. The x-axis represents glucocorticoid immunosuppression duration categories (days). Each gray bar indicates the count of patients in each duration group, and the black line represents the time ratio. The results indicate a decline in overall survival as immunosuppression duration increases, with smaller time ratios observed in higher dosage groups.

Our data also suggested a significantly negative impact on OS associated with the duration of immunosuppression administration (**Figure 4B**). Specifically, patients who received immunosuppressants for two to five days had an approximately 10-25% decrease in OS. Beyond five days, survival outcomes continue to worsen, approaching a 35% reduction in OS among patients receiving sISP for ≥7 days. Overall, indications for sISP use across different durations (**Figures S4B** and **S5B**) were diverse and multifactorial. Patients rarely received sISP for a single indication; instead, most were treated for a combination of reasons, including cancer symptom palliation, management of irAEs, and coexisting conditions such as pre-existing autoimmune diseases.

### Systemic immunosuppression among the irAE population

We identified 3,284 patients who developed irAEs in the MGBD cohort, among which 1,038 (31.6%) patients received sISP (**Table S21**), and 5,538 patients who developed irAEs in the TriNetX cohort, among which 1,299 (23.5%) patients received sISP (**Table S22**) within the [−3, 3] time window. Our results showed that irAE patients who received sISP within a year after ICI initiation had worse outcomes compared to irAE patients who did not receive sISP in that time frame (**Figure S6**). This observed negative effect was worst in patients receiving sISP nearest to ICI initiation and began to plateau after four months of ICI initiation.

## Discussion

As the primary form of immunosuppression in the ICI population, understanding the impact of systemic glucocorticoids on ICI outcomes is critical. To our knowledge, this is the largest study of the impact of sISP on survival among ICI recipients, analyzing ∼40,000 patients across three high-volume centers and a population-level cohort. In our study population, sISP use ranged from 17% to 37%, consistent with existing literature.^3,4,20^ Our comprehensive pan-cancer analyses enhance the generalizability of our findings and offer clinicians valuable insights on sISP use among ICI recipients.

In the MGBD cohort, patients who received sISP had a shorter median OS compared to those who did not receive sISP, which was validated in the TriNetX cohort. These results are consistent with existing literature, with one systematic review of 5,461 patients demonstrating increased mortality among non-small cell lung cancer patients requiring systemic corticosteroids after ICI initiation compared to non-exposed ICI recipients [pooled HR 1.62, p < 0.01].^21^ Another systematic review of mostly melanoma and lung cancer patients identified significantly worse OS and progression-free survival (PFS) among ICI recipients who received systemic corticosteroids for metastatic brain cancer compared to ICI recipients who did not.^22^ Additional investigations have elucidated the effect of timing, dosing, and indication of sISP on PFS and OS with variable results.^7–14^

This study comprehensively evaluates the effect of sISP timing on OS among ICI recipients, ranging from 12 months pre-ICI initiation to 12 months post-ICI initiation. Both univariate and multivariate analyses demonstrate that sISP use within this period is associated with a worse prognosis in the MGBD and TriNetX cohorts, which declines with closer timing between sISP administration and ICI initiation (**Figure 3**). Specifically, we demonstrate that patients receiving sISP within one month of ICI initiation exhibit the poorest survival [TR 0.49, 95%CI: 0.45-0.54; p<0.0001]. These results are consistent with prior studies, which examined the relationship between sISP at baseline 1 month prior to ICI and shortly after ICI initiation. Notable OS differences were identified among patients who received sISP between 60 days before ICI to 2 months post-ICI initiation.^10,14,21,23,24^ Similarly, Maslov et al demonstrated that patients receiving corticosteroids ≥2 months after ICI initiation had significantly improved OS compared with those who received corticosteroids <2 months after ICI initiation.^8^ However, these studies focused on limited timeframes around ICI start and largely did not explicitly examine the impact of pre-ICI sISP use. Though the detrimental effect of sISP on survival plateaued at 4 months post-ICI in both MGBD and TriNetX cohorts, patients receiving sISP remained at significantly increased risk of mortality through at least 12 months post-ICI initiation.

The only prior study to explicitly examine pre-ICI sISP leveraged a SEER-based population of 1,671 melanoma patients to pre-ICI sISP’s impact on survival.^23^ The authors found that patients with steroid exposure 1-3 months pre-ICI had worse survival compared to patients who were steroid naïve [HR 1.51, 95% CI: 1.01-2.27]. Consistent with our findings, patients with steroid exposure 1 month pre-ICI faced mortality risk [HR 2.26, 95% CI: 1.65-3.08]. However, no increased mortality risk was associated with steroid exposure 3 to 12 months pre-ICI. This was a key difference in our study findings which we suspect is due to the much smaller sample size of the prior investigation, limited the explanatory power of their analyses.

We further evaluated whether these trends were consistent among patients with irAEs and, therefore, more likely to require sISP. Since some types of irAEs (e.g., cutaneous and endocrine) have been previously associated with improved survival in the ICI population,^3,17,25^ it is unclear how the deleterious impact of sISP would manifest in this population, and results of prior studies have shown mixed results. One study found no difference in OS between patients receiving <10 mg prednisone and those receiving ≥10 mg prednisone daily for irAE management.^11^ However, this study did not delineate these differences in relation to sISP timing. In contrast, a large VA population-based study of 20,163 cancer patients identified decreased survival among patients receiving steroids for irAEs within 2 months post-ICI compared with later administrations.^24^ Our results demonstrate worse survival in patients with irAEs who received sISP compared to those with irAEs who did not throughout the 12 months post-ICI initiation (**Figure S6**). This effect was more prominent within the first 4 months of ICI initiation, as seen in the broader cohorts independent of sISP indication, suggesting sISP administration blunts the positive prognostic impact of irAE development. Thus, providers should exercise caution prescribing sISP near ICI initiation -even for the management of irAEs.

We suspect the trend in mortality risk pre- and post-ICI initiation, particularly the worse outcomes near ICI initiation, stems from dose-dependent inhibition of T-cell activation or proliferation in combination with the upregulation of PD-1 or CTLA-4 surface receptors on T-cells by glucocorticoids.^26,27^ These mechanisms facilitate tumor evasion of immune system surveillance and targeted response. Furthermore, the increased mortality seen with pre-ICI sISP administration is likely due to glucocorticoid effects lasting beyond initiation into the early post-ICI period. Mouse models have demonstrated that dexamethasone has a dose-dependent effect on PD-1 receptor upregulation that returned to baseline after 1-week.^28^ ICIs require weeks to months to be effective due to the gradual binding of ICI to PD-1, PD-L1, or CTLA-4 receptors. At 2-10mg/kg dosing of pembrolizumab, PD-L1 receptor saturation is expected within 3 weeks, and at ≥0.3mg/kg dosing of nivolumab, the saturation of PD-1 receptors with nivolumab is expected within 8 weeks.^29,30^ We suspect that once receptors reach saturation, glucocorticoid antagonism dampens, explaining the reduction in mortality risk for patients who received sISP after 3 months post-ICI.

We also identified dose- and duration-dependent impacts of sISP on survival. Most prior studies selected variable cut-off points to compare the effects of high-dose to low-dose sISP.^12,13,24^ In addition, glucocorticoid-naive patients were combined with low-dose sISP cohorts, obscuring the impact of low-dose sISP. We expand upon prior analyses by characterizing survival trends across a broad range of sISP doses. In the manually phenotyped MGBD cohort, any sISP dose ≥1 mg/day of prednisone equivalent reduced survival, with doses ≥75 mg/day linked to a nearly 40% decrease in survival time. Ricciuiti et al similarly found poorer survival in NSCLC patients receiving ≥10 mg of prednisone within 24 hours of ICI initiation, regardless of indication, compared to those receiving <10 mg.^13^ We identified key dose thresholds where mortality impact is greatest. Between 15 and 55 mg/day of prednisone equivalent, the mortality rate increases sharply. Above 65 mg/day, the rate of increase slows, plateauing at a nearly 40% reduction in survival time. Thus, providers should reduce the daily dose of sISP when feasible, targeting the dose limits discussed above.

Furthermore, Verheijden et al investigated peak dose corticosteroid usage via a pooled analysis of 6 clinical trials, finding that a peak median dose of 75 mg/day of prednisone equivalent was associated with increased mortality risk compared to lower doses.^7^ In contrast, cumulative dose was not associated with an impact on survival.^7^ Our findings confirm these results and expand on prior knowledge by providing detailed dose-specific impact of sISP on mortality among ICI recipients. Notably, an increasing proportion of patients received sISP for cancer-related indications at higher sISP doses (**Figure S4A**). Recent studies identified particularly poor survival among patients receiving sISP for cancer-related indications.^11–13^ However, our data suggest these outcomes may be mediated by higher administered sISP doses rather than greater cancer burden. Additionally, our study demonstrates that increasing duration of sISP exposure negatively impacts survival among ICI recipients, though this increased risk tends to stabilize at ≥7 days of exposure. Taken together, these findings suggest that limiting both sISP exposure, as well as duration and dose, may reduce the risk of mortality among ICI recipients.

The limitations of this study include its retrospective design and reliance on computational methods for data extraction. These methods are unable to obtain detailed data regarding medication adherence, which is particularly important for the TriNetX population where manual review of clinical notes is not possible. However, we validated our computational approaches through rigorous manual phenotyping of the MGBD cohort, demonstrating high concordance between manual and computational methods. Another limitation is that cancer-specific mortality data were not uniformly available across both large cohorts; potentially influencing the observed effect sizes. The median duration of follow-up, particularly in the TriNetX cohort, was relatively short, potentially leading to an underestimation of long-term effects. Furthermore, many patients were excluded from the MGBD and TriNetX cohorts prior to and during the propensity score matching process. Consequently, our findings are most generalizable to the final matched populations and may not fully represent the entirety of ICI recipients at these institutions or in the broader TriNetX network. Our investigation primarily focused on systemic glucocorticoids due to their predominance in the study population, and our ability to examine the independent impact of non-steroidal immunosuppression or specific steroid-sparing regimens (e.g., anti-TNFα, IL-6 inhibitors) was limited due to the lack of systematically available data on these agents, especially in the TriNetX cohort, and the inability to dissociate the effects of these agents from systemic corticosteroids which most patients use as first line immunosuppressive therapy. However, the effect of the association between sISP and mortality was larger in the MGBD cohort than TriNetX, where a greater proportion of the population also received non-glucocorticoid immunosuppression. Taken together, this suggests that non-steroidal immunosuppression is possibly less deleterious than glucocorticoids and may even be protective in some cases. Furthermore, extracting irAE severity is challenging using computational methods and may confound survival comparisons between sISP users and non-users, as the latter likely had less severe irAEs (E-value: 2.19).^31^ The handling of missing data for Hispanic ethnicity (Table S14-S17) suggests a potential Missing Not At Random (MNAR) scenario. The reasons for this missingness and its full impact on the results are unclear, warranting cautious interpretation of findings related to this variable and highlighting an area for further investigation. Additionally, while we accounted for guarantee-time bias in analyses, potential selection bias from estimating the correlation of sISP and survival is unavoidable, especially confounding by indication concerning early sISP administration, where patients requiring sISP, particularly early or for cancer-related symptoms, may have inherently more aggressive disease or a poorer performance status not fully captured by the measured covariates.

Nevertheless, our study leverages the largest cohort to date in evaluating the impact of sISP on OS across a pan-cancer population of ICI recipients, enhancing the generalizability of our findings. We further expand on previous studies by providing comprehensive information on the impact of sISP duration, dose, timing, and indication in the ICI population. These findings provide critical insights for clinicians and patients, suggesting that if sISP treatment is necessary, it should be administered with the lowest dose and shortest duration possible, avoiding administration near ICI initiation. Further studies are needed to explore the differential impact of non-steroidal or more targeted sISP therapies on ICI outcomes.

## Data Availability

All data produced in the present study are available upon reasonable request to the authors

## IRB approval status

The Mass General Brigham Institutional Review Board approved the study (Protocol #2020P002307). The study meets the criteria of secondary research, for which consent is not required.

## Data sharing

All relevant data are available from the corresponding author: Yevgeniy R. Semenov. All summary data supporting the findings of this study are available within the Article or its Appendix. The patient data generated at Massachusetts General Hospital, Brigham and Women’s Hospital, and Data-Farber Cancer Institute for this study can only be shared per specific institutional review board requirements. Upon a request to the corresponding author, a data-sharing agreement can be initiated following the institution specific guidelines. Patient data at the TriNetX network can be accessed following the TriNetX network guidelines.

## Contributors

Study conception and design: GW, NN, CL, SK, BY, AG, YRS.

Manual data collection: NN, GW, SK, BY, MA, BWL, WXC, AR, KT.

Data analysis and interpretation: GW, CL, NN, BY, AG, SK, YRS.

Manuscript writing: GW, NN, CL, AG, NRL, SK, YRS.

Study supervision: SGK, YRS.

Final approval of manuscript: All authors.

Accountable for all aspects of the work: All authors.

