## Supplemental tables for "Effect of systemic glucocorticoid immunosuppression timing, dose, and duration on overall survival among immune checkpoint inhibitor recipients: a retrospective multicohort study"

**Appendix**

| **1** | **Table S1**. Medication codes and names used to identify recipients of immune checkpoint inhibitors. |
| --- | --- |
| **2** | **Method S1**. Propensity score matching |
| **3** | **Table S2**. Definitions of variables included in this study |
| **4** | **Method S2**. The computational method for extracting systemic immunosuppression status. |
| **5** | **Method S3**. Manual chart review for immunosuppressive drugs. |
| **6** | **Table S3.** Patients by systemic immunosuppression category within the [-3, 3] window of the manual cohort. |
| **7** | **Figure S1**. Causal directed acyclic graph to identify potential confounding variables. |
| **8** | **Table S4**. ICD-9 and ICD-10 codes used to identify cancer type and stage. |
| **9** | **Table S5**. Medication names used to identify chemotherapy and targeted therapy. |
| **10** | **Table S6**. Systemic immunosuppressive drugs and categories. |
| **11** | **Method S4**. Sample size calculation. |
| **12** | **Table S7.** Conversion of non-steroidal immunosuppressive medication into Prednisone equivalent. |
| **13** | **Table S8**. Patient characteristics of the MGBD and TriNetX cohorts. |
| **14** | **Table S9.** Censoring proportion in every six-month interval within two-year follow-up duration in the MGBD and TriNetX cohorts. |
| **15** | **Table S10.** Patient characteristics of the TriNetX cohort stratified by glucocorticoid-induced sISP status. |
| **16** | **Figure S3**. Landmark survival analysis for the association of systemic immunosuppression with overall survival (Manual cohort). |
| **17** | **Table S11.** Concordance of extracting systemic immunosuppression data between the computational method and manual chart review in the [-3, 3] window. |
| **18** | **Figure S2**. Kaplan-Meier curves by time windows of systemic immunosuppression. |
| **19** | **Table S12.** Effect of systemic immunosuppression on overall survival using multivariable CoxPH modeling. |
| **20** | **Table S13.** Effect of systemic immunosuppression on overall survival using multivariable AFT modeling. |
| **21** | **Table S14.** The full AFT model for the association of sISP and overall survival with the [-1, 1] window (MGBD). |
| **22** | **Table S15**. The full AFT model for the association of sISP and overall survival with the [-3, 3] window (MGBD). |
| **23** | **Table S16**. The full AFT model for the association of sISP and overall survival with the [-1, 1] window (TriNetX). |
| **24** | **Table S17**. The full AFT model for the association of sISP and overall survival with the [-3, 3] window (TriNetX). |
| **25** | **Table S18**. Multivariable AFT modeling of systemic immunosuppression on overall survival (Manual cohort) |
| **26** | **Table S19**. Multivariable AFT modeling of systemic immunosuppression on overall survival (1:2 matching) |
| **27** | **Table S20.** Indications for systemic immunosuppression (Manual). |
| **28** | **Figure S4.** Reasons for immunosuppression with different dosage and duration categories (Manual cohort). |
| **29** | **Figure S5:** Reasons for immunosuppression across different dosage and duration categories in patients exclusively treated with glucocorticoids (Manual Cohort) |
| **30** | **Table S21**. Characteristics of irAE patients stratified by gsISP status (MGBD). |
| **31** | **Table S22**. Characteristics of irAE patients stratified by gsISP status (TriNetX). |
| **32** | **Figure S6**. Landmark survival analyses for the association between systemic immunosuppression and overall survival among irAE patients. |
| **33** | **Figure S7.** Reasons for Immunosuppression Across Different Dosage and Duration Categories in Patients Exclusively Treated with Glucocorticoids (Manual Cohort) |

**Table S1.** Medication codes and names used to identify recipients of immune checkpoint inhibitors.

| **ICI Type** | **Code Type** | **Code** | **Medication Name** |
| --- | --- | --- | --- |
| CTLA-4 | HCPCS | J9228 | Ipilimumab |
| CTLA-4 | RxNorm | 1094833 | Ipilimumab |
| PD-1 | RxNorm | 1597876 | Nivolumab |
| PD-1 | HCPCS | J9299 | Nivolumab |
| PD-1 | HCPCS | J9271 | Pembrolizumab |
| PD-1 | RxNorm | 1547545 | Pembrolizumab |
| PD-1 | HCPCS | J9119 | Cemiplimab |
| PD-1 | RxNorm | 2058826 | Cemiplimab |
| PD-L1 | HCPCS | J9023 | Avelumab |
| PD-L1 | RxNorm | 1875534 | Avelumab |
| PD-L1 | HCPCS | J9022 | Atezolizumab |
| PD-L1 | RxNorm | 1792776 | Atezolizumab |
| PD-L1 | ICD-10-PCS | XW043D6 | Atezolizumab |
| PD-L1 | ICD-10-PCS | XW033D6 | Atezolizumab |
| PD-L1 | HCPCS | J9173 | Durvalumab |
| PD-L1 | RxNorm | 1919503 | Durvalumab |

CTLA-4: cytotoxic T-lymphocyte-associated protein 4; PD-1: programmed death receptor-1; PD-L1: programmed death-ligand 1.

**Method S1.** Propensity score matching

The TriNetX cohort (N = 26,172) was identified by a 2:1 propensity score matching to the MGBD cohort (N = 13,086) by the baseline characteristics (self-reported sex, race, ethnicity, age at ICI, Charlson comorbidity index, cancer type, cancer stage, ICI therapy type, and pre-ICI treatment) as well as the year of ICI initiation (Table S2 for definitions). The goal of this matching is to ensure a basic level of similarity between the two cohorts.

We used the "matchit" function of the MatchIt package (version 4.3.3) in R. The default parameter values of the function were used. By default, the propensity score was estimated with logistic regression, and the nearest neighbor matching on the propensity score with no caliper and no replacement was conducted.

**Table S2.** Definitions of variables included in this study.

| **Index** | **Variable** | **Definition** |
| --- | --- | --- |
| 1 | Sex | Self-reported biological sex extracted from electronic health records. |
| 2 | Race | Self-reported race from electronic health records. For missing race, "Unavailable" was assigned to the variable. |
| 3 | Ethnicity | Self-reported ethnicity from electronic health records. For missing ethnicity, "Unavailable" was assigned to the variable. |
| 4 | Age at initiation of immune checkpoint inhibitor therapy  (Age at ICI) | Age on the date of first immune checkpoint inhibitor administration.  The variable was grouped based on the impact on overall survival and sample size: <=60, 61–70, 71–80, and >80. |
| 5 | Charlson comorbidity index (CCI) | Computed using patient diagnostic codes from the International Classification of Diseases Versions 9 and 10 before initiation of immune checkpoint inhibitor therapy. |
| 6 | Cancer type | Extracted using diagnostic codes specified in Table S4. |
| 7 | Cancer stage | Estimated using secondary malignancy diagnostic codes (Table S4):  "metastatic" if a patient had secondary cancer in distant sites;  "locoregional" if a patient had no secondary cancer diagnoses or had secondary cancer limited to lymph nodes. |
| 8 | Immune checkpoint inhibitor type  (ICI type) | Immune checkpoint inhibitor therapy type specified in Table S1. |
| 9 | Number of immune checkpoint inhibitor therapy cycles  (ICI cycles) | Immune checkpoint inhibitor therapy is commonly administered on a varied schedule (often every three weeks), spanning several weeks to potentially up to two years. This study included the number of cycles of immune checkpoint inhibitor therapy a patient received in our survival models with an upper limit of five to mitigate the potential bias from frequent healthcare utilization. As a result, the possible values of this variable are: 1, 2, 3, 4, and >= 5. |
| 10 | Pre-immune checkpoint inhibitor treatment  (Pre-ICI treatment) | Cytotoxic chemotherapy and targeted anti-neoplastic therapy were identified by medication names (Tables S5). Pre-ICI treatment was categorized as:  "conventional chemotherapy" if a patient received cytotoxic chemotherapy (and/or targeted therapy) before immune checkpoint inhibitor therapy;  "targeted therapy" if a patient received targeted therapy alone before initiation of immune checkpoint inhibitor therapy;  "none" otherwise. |
| 11 | Concurrent non-immune checkpoint inhibitor treatment  (Concurrent non-ICI treatment) | Concurrent other treatment was categorized as follows:  "conventional chemotherapy" if a patient received cytotoxic chemotherapy (and/or targeted therapy) within two years after initiation of immune checkpoint inhibitor therapy;  "targeted therapy" if a patient received targeted therapy alone within two years after initiation of immune checkpoint inhibitor therapy;  "none" otherwise. |
| 12 | Non-immune checkpoint inhibitor treatment  (Non-ICI treatment) | In landmark analyses, we combined "pre-immune checkpoint inhibitor treatment" and "concurrent non-immune checkpoint inhibitor treatment" as "non-immune checkpoint inhibitor treatment" due to the potential correlation between them:  "conventional chemotherapy" if a patient received cytotoxic chemotherapy (and/or targeted therapy) between three months before immune checkpoint inhibitor therapy initiation to the landmark time;  "targeted therapy" if a patient received targeted therapy alone between three months before immune checkpoint inhibitor initiation to the landmark time;  "none" otherwise. |
| 13 | Year of immune checkpoint inhibitor therapy initiation  (Year of ICI) | The year of immune checkpoint inhibitor therapy initiation |
| 14 | Duration of immune checkpoint inhibitor, days  (Duration of ICI) | Days from initiation of immune checkpoint inhibitor therapy to last administration |
| 15 | sISP reason | This variable considered the treatment reason of systemic immunosuppressants (sISP):  "irAEs": includes patients who received sISP for managing irAEs and/or other conditions.  "Cancer palliation": includes patients who received sISP for cancer palliation and/or other conditions, except irAEs.  "Pre-existing condition": includes patients who received sISP for managing pre-existing and/or other conditions, except irAEs and cancer palliation.  "Other": includes patients who received sISP for conditions not mentioned above, such as transplantation, allergies, infection. |
| 16 | ISP dosage | Dosage was calculated as the total cumulative dose over the total number of days on immunosuppressants, stratifying patients along dosage thresholds 1+, 5+, 10+, 20+, 40+, 80+, 120+, 200+, 300+, 400+, 500+, 600+, 700+, 800+, 900+, and 1000+ mg/day. |
| 17 | ISP duration | The total number of days was used to determine the duration of calculated as the total cumulative dose over the total number of days on systemic immunosuppression use, stratifying using 1+, 2+, 3+, 5+, 7+, 14+, 21+, 28+, 35+ day thresholds. |
| 18 | Computational sISP | This variable was defined based on the systemic glucocorticoid use in the inpatient and outpatient settings relative to immune checkpoint inhibitor therapy initiation:  "Yes", if  (1) a patient was treated with systemic glucocorticoids in the inpatient setting for twice, or  (2) a patient was treated with prednisone >= 10mg twice.  "No" otherwise.  Method S2 presents the details for extracting this variable. |
| 19 | Mortality status at end of follow-up | "Dead" if death was reported for a patient;  "alive" otherwise. |
| 20 | Duration of follow-up, days | Days from initiation of immune checkpoint inhibitor therapy to date of death or last follow-up. |

**Method S2.** The computational method for extracting systemic immunosuppression status.

Guided by our manual chart review for immunosuppression data extraction (Method S3), we established an approach to identify systemic immunosuppression (sISP) use computationally. Here, we focused on glucocorticoid-induced sISP since we observed that among manually reviewed patients who received sISP, only one did not receive glucocorticoids (Table S3). Specifically, we preprocessed the data for both cohorts as follows:

- Extracted glucocorticoid administration records (Table S6) and encounter information from both databases.
- Excluded records of inhaled medications, ophthalmic solutions, drops, and creams.
- Categorized patients as "inpatient", if a patient was treated with systemic glucocorticoids in the inpatient setting; "outpatient", if a patient was treated with systemic glucocorticoids in the outpatient setting during the period; "no" otherwise.

The "Computational sISP" variable was defined based on the systemic glucocorticoid use in the inpatient and outpatient settings relative to immune checkpoint inhibitor (ICI) therapy initiation:

"Yes", if

(1) a patient was treated with systemic glucocorticoids in the inpatient setting for twice, or

(2) a patient was treated with prednisone >= 10mg twice.

"No" otherwise.

To evaluate the accuracy of this computational sISP identification, we compared this variable to the glucocorticoid sISP status (Yes or No) extracted using manual chart review.

**Method S3.** Manual chart review for immunosuppressive drugs.

A comprehensive list of immunosuppressive drugs (Table S6) was used to extract immunosuppressant (ISP) administration records from our multi-institutional electronic medical data warehouse for a subset of 1,128 patients treated by immune checkpoint inhibitor (ICI) therapy at MGBD. The evaluation timeframe was from 12 months before ICI initiation until one year after ICI initiation or until the date of last follow-up.

Records of inhaled medications, ophthalmic solutions, drops, and creams were excluded. Patients without any records were considered as "no systemic ISP". Manual chart review was conducted for patients with records of ISP use during the evaluation timeframe. Inpatient and outpatient immunosuppressant orders were manually reviewed separately and then merged. Patients who were prescribed medications were assumed to have taken the medication. All orders with the same medication, dose, and overlapping were chart reviewed for duplicates. All systemic glucocorticoid medications were converted to oral prednisone equivalent.^4,5^

Patient cumulative dosage and total days of systemic ISP (sISP) exposure were manually calculated. Dates and categories (Systemic Glucocorticoids, Biologics, and Oral Non-Steroidal Systemic) of sISP were recorded. Systemic ISP was defined based on immunosuppressant administration guidelines. For glucocorticoids, which represented the majority of ISP in our cohort, patients receiving 10mg prednisone equivalent daily for at least seven days continuously were considered as patients with sISP. For non-steroidal systemic therapy, receiving one week of treatment per prescription for at least two prescriptions at least one month apart was considered sISP. For methotrexate specifically, once weekly for twelve weeks was considered sISP. Infliximab, vedolizumab, and IVIG use were considered sISP if given once monthly. Rituximab was considered sISP if given once in six months. Bevacizumab was considered sISP if given once in two weeks. Adalimumab and tocilizumab were considered sISP if given every two weeks. Etanercept was considered sISP if given once weekly.

Reasons for sISP treatments were manually extracted from which patients were classified into two categories: 1) immune-related adverse events (irAEs), if a patient received systemic ISP for managing irAEs with or without other reasons; 2) other, if a patient received systemic ISP for other reasons, including cancer palliation, preexisting conditions, and other reasons.

In summary, we included four variables: sISP status (Yes or No); sISP reason (irAEs or Other), sISP total dosage, and sISP categories (Systemic Glucocorticoids, Biologics, and Oral Non-Steroidal Systemic) within one year before ICI initiation until one year after ICI start in our study.

**Table S3.** Patients by systemic immunosuppression category within the [-3, 3] window of the manual cohort.

| **Systemic immunosuppression category** | **Number of Patients Receiving sISP**^1^  **(N = 415)** |
| --- | --- |
| Systemic Glucocorticoid and/or others | 414 (99.8%) |
| Biologic and/or others | 12 (2.9%) |
| Oral Systemic Non-Steroidal and/or others | 11 (2.7%) |

^1^Out of the 1,128 patients of the manual cohort, 415 (36.8%) patients received systemic immunosuppressants within the [-3, 3] window centered by the initiation of immune checkpoint inhibitor therapy (time zero).

sISP: systemic immunosuppression.

**Figure S1.** Causal directed acyclic graph to identify potential confounding variables.

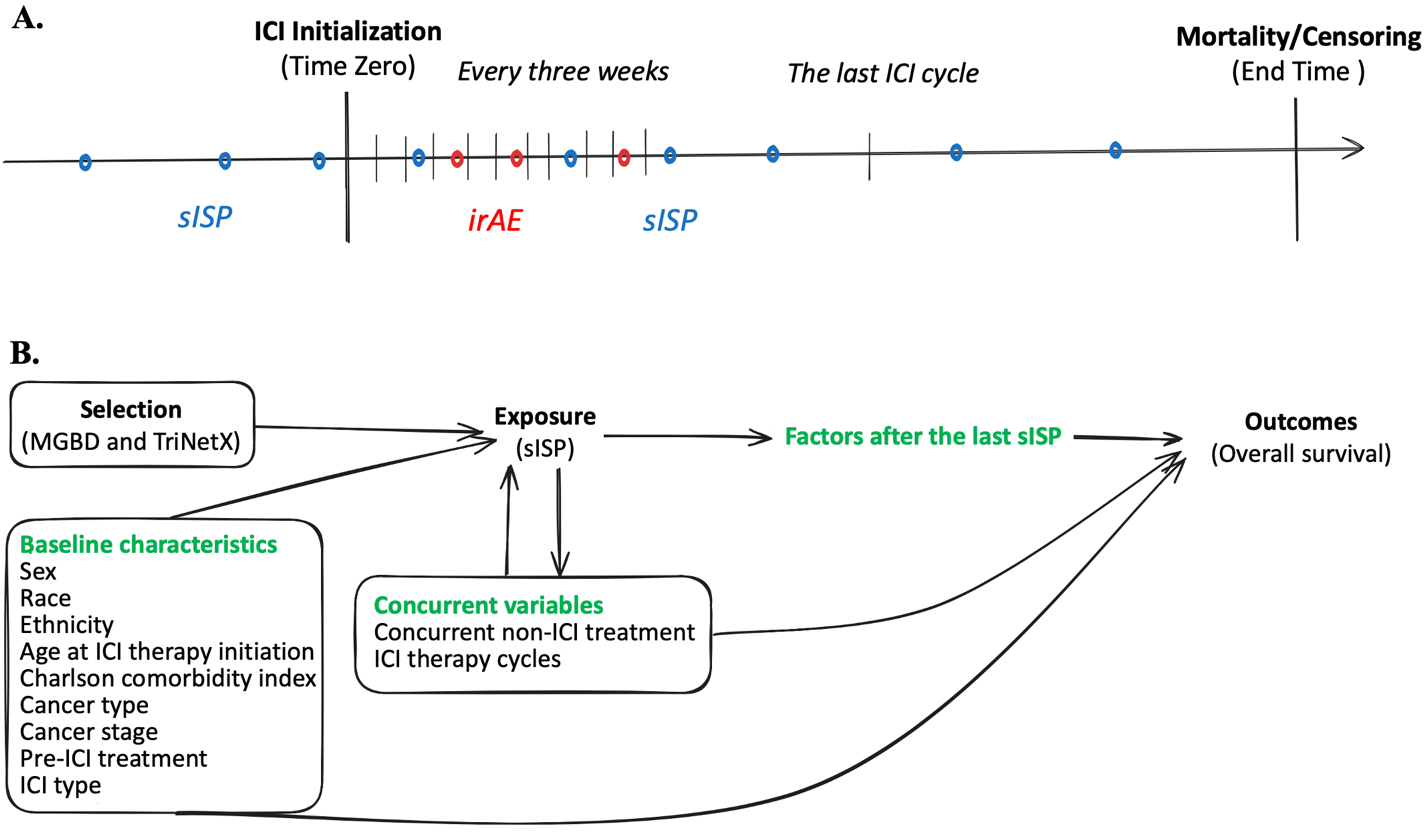

**Legend:**

**A.** The study timeline. The time zero, or the time origin, for this study is the start date of immune checkpoint inhibitor (ICI) therapy. ICI therapy is commonly administered on a varied schedule, often spanning several weeks to potentially up to two years, contingent upon clinical considerations. The focus of this study is on systemic immunosuppression (sISP), in particular from glucocorticoids. We considered glucocorticoid sISP within 1.5 years following the initiation of ICI therapy. The end time of this study is the date of death or the date of censoring. For patients who did not have known mortality information, their last encounter was considered as their censoring date. ICI recipients may develop immune-related adverse events (irAEs). We conducted experiments among irAE patients.

**B.** Causal directed acyclic graph (cDAG). We grouped variables into three categories: baseline characteristics (measured at the time of ICI initiation), concurrent variables (occurred during ICI therapy before the last glucocorticoid sISP administration), and factors after the last glucocorticoid sISP administration. We considered baseline and concurrent variables as potential confounders. (In causal inference, factors after exposure cannot be confounders.) Our survival models were adjusted by baseline characteristics, non-ICI treatment (stratified by conventional chemotherapy and targeted therapy), and ICI therapy cycles. Table S2 provides definitions of the variables included in this study.

**Table S4.** ICD-9 and ICD-10 codes used to identify cancer type and stage.

| **Cancer Type** | **ICD-10** | **ICD-9** |
| --- | --- | --- |
| Melanoma | C43 | 172 |
| Thoracic | C30-C39 | 160-165 |
| Digestive | C15-C26 | 150-159 |
| Female genital | C51-C58 | 179-184 |
| Male genital or urinary | C60-C68 | 185-189 |
| Lymphoid or haematopoietic | C81-C96 | 200-209 |
| Breast | C50 | 174-175 |
| Brain, nervous, or eye | C69-C72 | 190-192 |
| Oral, lip, pharynx | C00-C14 | 140-149 |
| Non-melanoma skin malignancy | C44, C4A | 173 |
| Other | Start with 'C' but do not fall into the categories above | 140-209 but do not fall into the categories above |
| Secondary cancer: lymph nodes | C77 | 196 |
| Secondary cancer: distant sites | C78-C79 | 197-198 |

ICD-9: International Classification of Diseases version 9.

ICD-10: International Classification of Diseases version 10.

**Table S5.** Medication names used to identify chemotherapy and targeted therapy.

| **Therapy Type** | **Medication Name** |
| --- | --- |
| Cytotoxic chemotherapy | Daunorubicin, Cytarabine, Arsenic, Asparaginase, Azacitidine, Bendamustine, Busulfan, Capecitabine, Carboplatin, Carmustine, Chlorambucil, Cisplatin, Caldribine, Clofarabine, Cyclophosphamide, Dacarbazine, Decitabine, Degarelix, Docetaxel, Eribulin Mesylate, Etoposide, Floxiridine, Fludarabine, Fluorouracil, Gemcitabine, Goserelin, Histrelin, Hydroxyurea, Ifosfamide, Interferon, Irinotecan, Ixabepilone, Lanreotide, Leucovorin, Lomustine, Medroxyprogesterone, Megestrol, Melphalan, Mercaptopurine, Mesna, Methotrexate, Mitoxantrone, Nelarabine, Octreotide, Omacetaxine Mepesuccinate, Oxaliplatin, Paclitaxel, Pegaspargase, Pemetrexed, Pentostatin, Procarbazine, Samarium SM 153 Lexidronam, Temozolomide, Teniposide, Thiotepa, Topotecan, Trabectedin, Vinblastine, Vinorelbine, Doxorubicin, Idarubicin, Epirubicin, Bleomycin, Cabazitaxel, Dactinomycin, Mitomycin, Streptozocin |
| Targeted anti-neoplastic  therapy | Trastuzumab, Aflibercept, Alemtuzumab, Anastrozole, Belimumab, Belinostat, Bevacizumab, Blinatumomab, Bortezomib, Brentuximab Vedotin, Carfilzomib, Cetuximab, Copanlisib, Denosumab, Elotuzumab, Everolimus, Exemestane, Fulvestrant, Gefitinib, Gemtuzumab Ozogamicin, Imatinib Mesylate, Inotuzumab Ozogamicin, Mogamulizumab-kpkc, Moxetumomab Pasudotox-tdfk, Natalizumab, Necitumumab, Obinutuzumab, Ofatumumab, Olaratumab, Panitumumab, Pertuzumab, Pralatrexate, Radium, Ramucirumab, Rituximab, Romidepsin, Siltuximab, Tagraxofusp, Talazoparib, Tamoxifen, Temsirolimus, Valrubicin, Ziv-Aflibercept, Bicalutamide, Floxuridine, Flutamide, Ketoconazole, Lanreotide, Lenalidomide, Levoleucovorin, Pomalidomide, Porfimer Sodium, Propylthiouracil, Thalidomide, Trifluridine, Vincristine, Abemaciclib, Abiraterone, Acalabrutinib, Afatinib, Alectinib, Alpelisib, Apalutamide, Axitinib, Binimetinib, Bosutinib, Brigatinib, Cabozantinib S-Malate, Ceritinib, Cobimetinib, Crizotinib, Dabrafenib, Dacomitinib, Daratumumab, Darolutamide, Dasatinib, Dinutuximab, Duvelisib, Elotuzumab, Enasidenib, Encorafenib, Enfortumab Vedotin, Entrectinib, Enzalutamide, Erdafitinib, Erlotinib, Fam-Trastuzumab Deruxtecan-nxki, Fedratinib Hydrochloride, Fulvestrant, Gilteritinib, Glasdegib, Ibrutinib, Idelalisib, Isatuximab, Isotretinoin, Ivosidenib, Ixazomib, Lapatinib, Larotrectinib, Lenvatinib, Letrozole, Lorlatinib, Neratinib, Nilotinib, Niraparib, Obinutuzumab, Olaparib, Olaratumab, Osimertinib, Palbociclib, Panobinostat, Pazopanib, Pexidartinib, Polatuzumab Vedotin, Ponatinib, Regorafenib, Ribociclib, Rucaparib, Ruxolitinib, Midostaurin, Selinexor, Sonidegib, Sorafenib, Sunitinib, Talazoparib Tosykate, Tazemetostat, Toremifene, Trametinib, Tretinoin, Vandetanib, Vemurafenib, Venetoclax, Vismodegib, Vorinostat, Zanubrutinib, Ziv-Aflibercept |

**Table S6.** Systemic immunosuppressive drugs and categories.

| **Systemic Glucocorticoid** | **Biologics** | **Oral Non-Steroidal Systemic** |
| --- | --- | --- |
| Prednisone | Etanercept | Sirolimus |
| Methylprednisolone | Abatacept | Tacrolimus |
| Hydrocortisone | Infliximab | Tofacitinib |
| Budesonide | Rituximab | Leflunomide |
| Cortisone | Adalimumab | Methotrexate |
| Prednisolone | Certolizumab | Azathioprine |
| Betamethasone | Golimumab | Everolimus |
| Dexamethasone | Tocilizumab | Cyclosporine |
| Corticotropin | Vedolizumab | Cyclophosphamide |
|  | Anakinra | Mycophenolate |
|  | Belimumab | Paclitaxel |
|  | Ixekizumab | Sulfasalazine |
|  | Natalizumab | Mesalamine |
|  | Secukinumab | Lenalidomide |
|  | Ustekinumab | Apremilast |
|  | Brodalumab | Penicillamine |
|  | Guselkumab | Balsalazide |
|  | Sarilumab | Chlorambucil |
|  |  | Hydroxyurea |
|  |  | Mepacrine |
|  |  | Mycophenolate Mofetil |
|  |  | Baricitinib |
|  |  | Olsalazine |
|  |  | Pimecrolimus |
|  |  | Mercaptopurine |
|  |  | Upadacitinib |
|  |  | Thalidomide |
|  |  | Temsirolimus |
|  |  | Thioguanine |
|  |  | Dimethyl Fumarate |

**Method S4.** Sample size calculation.

Sample size determination was conducted via a simulation-based power calculation. Iterative simulations estimated the required sample size (n) to achieve a target power of 80% (α= 0.05). Simulations began at n = 200, with stepwise increments of 100 (reduced to 20 near the target power > 75%) up to a maximum of n = 10,000. For every n value, 1000 random cohorts were subset from MGDB and TriNetX cohorts, and accelerated failure time (AFT) models with log-normal distributions were adjusted by sex, age at ICI initiation, race, ethnicity, cancer type, cancer stage, Charlson Comorbidity index, ICI type, and ICI cycles. Power was defined as the proportion of simulations yielding p < 0.05 for the primary predictor, sISP status. The iterative process terminated when the target power was reached.

**Table S7.** Conversion of non-steroidal immunosuppressive medication into Prednisone equivalent.

| **Corticosteroid** | **Equivalent Dose (mg) to 5 mg Prednisone** | **Duration of Action** |
| --- | --- | --- |
| **Prednisone** | 5 mg | Intermediate |
| **Prednisolone** | 5 mg | Intermediate |
| **Methylprednisolone** | 4 mg | Intermediate |
| **Dexamethasone** | 0.75 mg | Long |
| **Hydrocortisone** | 20 mg | Short |
| **Cortisone** | 25 mg | Short |
| **Betamethasone** | 0.75 mg | Long |
| **Triamcinolone** | 4 mg | Intermediate |

**Table S8.** Patient characteristics of the MGBD and TriNetX cohorts.

| **Characteristics**^1^ | **MGBD**  (N=13,086) | **TriNetX**  (N=26,172) |
| --- | --- | --- |
| **Sex** |  |  |
| Female | 6,072 (46.4%) | 11,671 (44.6%) |
| Male | 7,014 (53.6%) | 14,501 (55.4%) |
| **Race** |  |  |
| White | 11,791 (90.1%) | 23,534 (89.9%) |
| Black or African American | 364 (2.8%) | 746 (2.9%) |
| Asian | 430 (3.3%) | 787 (3.0%) |
| Other/Unavailable | 501 (3.8%) | 1,105 (4.2%) |
| **Ethnicity** |  |  |
| Not Hispanic | 11,896 (90.9%) | 23,663 (90.4%) |
| Hispanic | 366 (2.8%) | 849 (3.2%) |
| Unavailable | 824 (6.3%) | 1,660 (6.3%) |
| **Age at ICI Initiation (years)** |  |  |
| ≤60 | 4,169 (31.9%) | 8,343 (31.9%) |
| 61–70 | 4,157 (31.8%) | 8,456 (32.3%) |
| 71–80 | 3,399 (26.0%) | 7,056 (27.0%) |
| >80 | 1,361 (10.4%) | 2,317 (8.9%) |
| **Charlson Comorbidity Index** |  |  |
| 0 | 77 (0.6%) | 192 (0.7%) |
| 1–2 | 1,661 (12.7%) | 3,407 (13.0%) |
| 3–4 | 927 (7.1%) | 1,995 (7.6%) |
| ≥5 | 10,421 (79.6%) | 20,578 (78.6%) |
| **Cancer Type** |  |  |
| Thoracic | 3,325 (25.4%) | 7,035 (26.9%) |
| Male Genital/Urinary | 1,760 (13.4%) | 3,813 (14.6%) |
| Digestive | 1,626 (12.4%) | 3,258 (12.4%) |
| Melanoma | 1,318 (10.1%) | 2,833 (10.8%) |
| Other Skin Malignancy | 697 (5.3%) | 1,337 (5.1%) |
| Breast | 896 (6.8%) | 1,366 (5.2%) |
| Lymphoid/Hematopoietic | 713 (5.4%) | 1,267 (4.8%) |
| Female Genital | 636 (4.9%) | 1,153 (4.4%) |
| Brain/Nervous/Eye | 552 (4.2%) | 804 (3.1%) |
| Oral/Lip/Pharynx | 488 (3.7%) | 1,026 (3.9%) |
| Other | 1,075 (8.2%) | 2,280 (8.7%) |
| **Cancer Stage** |  |  |
| Distant | 10,252 (78.3%) | 20,234 (77.3%) |
| Locoregional | 2,834 (21.7%) | 5,938 (22.7%) |
| **Pre-ICI Treatment** |  |  |
| Conventional Chemotherapy | 4846 (37.0%) | 10264 (39.22%) |
| Targeted Therapy | 1499 (11.5%) | 2699 (10.31%) |
| None | 6741 (51.5%) | 13209 (50.47%) |
| **ICI Type** |  |  |
| PD-1 | 9,905 (75.7%) | 19,551 (76.2%) |
| PD-L1 | 1,836 (14.0%) | 3,592 (13.7%) |
| CTLA-4 | 142 (1.1%) | 241 (0.9%) |
| Combination^2^ | 1,203 (9.2%) | 2,388 (9.1%) |
| **Year of ICI Initiation** |  |  |
| Before 2017 | 2,147 (16.4%) | 4,051 (15.5%) |
| 2017 | 2,042 (15.6%) | 3,924 (15.0%) |
| 2018 | 2,355 (18.0%) | 4,689 (17.9%) |
| 2019 | 2,386 (18.2%) | 4,887 (18.7%) |
| 2020 | 2,212 (16.9%) | 4,603 (17.6%) |
| 2021 | 1,944 (14.9%) | 4,018 (15.4%) |
| **Duration of ICI (days)** |  |  |
| Median [IQR] | 106 [29, 316] | 92 [22, 279] |
| **Mortality status** |  |  |
| Alive | 5,728 (43.8%) | 16,151 (61.7%) |
| Dead | 7,358 (56.2%) | 10,021 (38.3%) |
| **Duration of Follow-up (days)** |  |  |
| Median [IQR] | 317 [113, 712] | 249 [91, 616] |

^1^ Definitions of variables are provided in Table S2.

^2^ Combination therapy of CTLA-4 and PD-1/PD-L1.

ICI: immune checkpoint inhibitor; IQR: interquartile range; PD-1: programmed death receptor-1; PD-L1: programmed death-ligand 1; CTLA-4: cytotoxic T-lymphocyte-associated protein 4; IQR: interquartile range.

**Table S9.** Censoring proportion in every six-month interval within two-year follow-up duration in the MGBD and TriNetX cohorts.

|  |  | **6 months** | **12 months** | **18 months** | **24 months** |
| --- | --- | --- | --- | --- | --- |
| **MGBD**  (N=13,086) | **Death** | 3,269 (25.0%) | 1,642 (12.5%) | 978 (7.5%) | 520 (4.0%) |
|  | **Loss of follow-up** | 1,218 (9.3%) | 1,030 (7.9%) | 708 (5.4%) | 545 (4.2%) |
|  | **Cumulative** | 4,487 (34.3%) | 7,159 (54.7%) | 8,845 (67.6%) | 9,910 (75.7%) |
| **TriNetX**  (N=26,172) | **Death** | 5,035 (19.2%) | 2,235 (8.5%) | 1,148 (4.4%) | 679 (2.6%) |
|  | **Loss of follow-up** | 5,707 (21.8%) | 2,948 (11.3%) | 1,704 (6.5%) | 1,375 (5.3%) |
|  | **Cumulative** | 10,742 (41.0%) | 15925 (60.8%) | 18,777 (71.7%) | 20,831 (79.6%) |

Columns present the patients (percentage %) of the population that died or were lost to follow-up in each six-month period across the MGBD and TriNetX populations. Cumulative censoring proportions are presented in the final row for each cohort.

**Table S10.** Patient characteristics of the TriNetX cohort stratified by glucocorticoid-induced sISP status.

| **Characteristics**^1^ | **sISP**  (N=4,526) | **No sISP**  (N=21,646) | **P-value** |
| --- | --- | --- | --- |
| **Sex** |  |  |  |
| Female | 2,012 (44.5%) | 9,659 (44.6%) | 0.85 |
| Male | 2,514 (55.5%) | 11,987 (55.4%) |  |
| **Race** |  |  |  |
| White | 4,130 (91.3%) | 19,404 (89.6%) | 0.0041 |
| Black or African American | 126 (2.8%) | 620 (2.9%) |  |
| Asian | 113 (2.5%) | 674 (3.1%) |  |
| Other/Unavailable | 157 (3.5%) | 948 (4.4%) |  |
| **Ethnicity** |  |  |  |
| Not Hispanic | 4,193 (92.6%) | 19,470 (89.9%) | < 0.0001 |
| Hispanic | 132 (2.9%) | 717 (3.3%) |  |
| Unavailable | 201 (4.4%) | 1,459 (6.7%) |  |
| **Age at ICI Initiation (years)** |  |  |  |
| ≤60 | 1,563 (34.5%) | 6,780 (31.3%) | < 0.0001 |
| 61–70 | 1,464 (32.3%) | 6,992 (32.3%) |  |
| 71–80 | 1,114 (24.6%) | 5,942 (27.5%) |  |
| >80 | 385 (8.5%) | 1,932 (8.9%) |  |
| **Charlson Comorbidity Index** |  |  |  |
| 0 | 36 (0.8%) | 156 (0.7%) | 0.0022 |
| 1–2 | 556 (12.3%) | 2,851 (13.2%) |  |
| 3–4 | 403 (8.9%) | 1,592 (7.4%) |  |
| ≥5 | 3,531 (78.0%) | 17,047 (78.8%) |  |
| **Cancer Type** |  |  |  |
| Thoracic | 1,441 (31.8%) | 5,594 (25.8%) | < 0.0001 |
| Male Genital/Urinary | 562 (12.4%) | 3,251 (15.0%) |  |
| Digestive | 461 (10.2%) | 2,797 (12.9%) |  |
| Melanoma | 519 (11.5%) | 2,314 (10.7%) |  |
| Other Skin Malignancy | 166 (3.7%) | 1,171 (5.4%) |  |
| Breast | 196 (4.3%) | 1,170 (5.4%) |  |
| Lymphoid/Hematopoietic | 265 (5.9%) | 1,002 (4.6%) |  |
| Female Genital | 155 (3.4%) | 998 (4.6%) |  |
| Brain/Nervous/Eye | 222 (4.9%) | 582 (2.7%) |  |
| Oral/Lip/Pharynx | 151 (3.3%) | 875 (4.0%) |  |
| Other | 388 (8.6%) | 1,892 (8.7%) |  |
| **Cancer Stage** |  |  |  |
| Locoregional | 1,064 (23.5%) | 4,874 (22.5%) | 0.15 |
| Distant | 3,462 (76.5%) | 16,772 (77.5%) |  |
| **Non-ICI Treatment** |  |  |  |
| Conventional Chemotherapy | 2,105 (46.5%) | 9,410 (43.5%) | < 0.0001 |
| Targeted Therapy | 401 (8.9%) | 2,755 (12.7%) |  |
| None | 2,020 (44.6%) | 9,481 (43.8%) |  |
| **ICI Type** |  |  |  |
| PD-1 | 3,284 (72.6%) | 16,667 (77.0%) | < 0.0001 |
| PD-L1 | 579 (12.8%) | 3,013 (13.9%) |  |
| CTLA-4 | 66 (1.5%) | 175 (0.8%) |  |
| Combination^2^ | 597 (13.2%) | 1,791 (8.3%) |  |
| **ICI Treatment Cycles** |  |  |  |
| 1 | 1,244 (27.5%) | 4,424 (20.4%) | < 0.0001 |
| 2 | 849 (18.8%) | 2,693 (12.4%) |  |
| 3 | 725 (16.0%) | 2,844 (13.1%) |  |
| 4 | 750 (16.6%) | 4,201 (19.4%) |  |
| ≥5 | 958 (21.2%) | 7,484 (34.6%) |  |
| **Mortality status** |  |  |  |
| Alive | 2,556 (56.5%) | 13,595 (62.8%) | < 0.0001 |
| Dead | 1,970 (43.5%) | 8,051 (37.2%) |  |
| **Duration of Follow-up (days)** |  |  |  |
| Median [IQR] | 125 [50, 336] | 285 [107, 663] | < 0.0001 |

^1^ Definitions of variables are provided in Table S2. Systemic immunosuppression (sISP): Patients treated with systemic glucocorticoids within three months before or after initiation of ICI therapy ([-3, 3]) were considered in the sISP group; otherwise in the no sISP group.

^2^ Combination therapy of CTLA-4 and PD-1/PD-L1.

ICI: immune checkpoint inhibitor; sISP: systemic immunosuppression; IQR: interquartile range; PD-1: programmed death receptor-1; PD-L1: programmed death-ligand 1; CTLA-4: cytotoxic T-lymphocyte-associated protein 4; IQR: interquartile range.

**Table S11.** Concordance of extracting systemic immunosuppression data between the computational method and manual chart review in the [-3, 3] window.

| **Concordance** | **PPV** | **NPV** |
| --- | --- | --- |
| 0.83  95% CI [0.80, 0.85] | 0.85  95% CI [0.81, 0.89] | 0.82  95% CI [0.79, 0.84] |

PPV: Positive Predictive Value; NPV: Negative Predictive Value.

**Figure S2.** Kaplan-Meier curves by time windows of systemic immunosuppression.

**A.** Manual

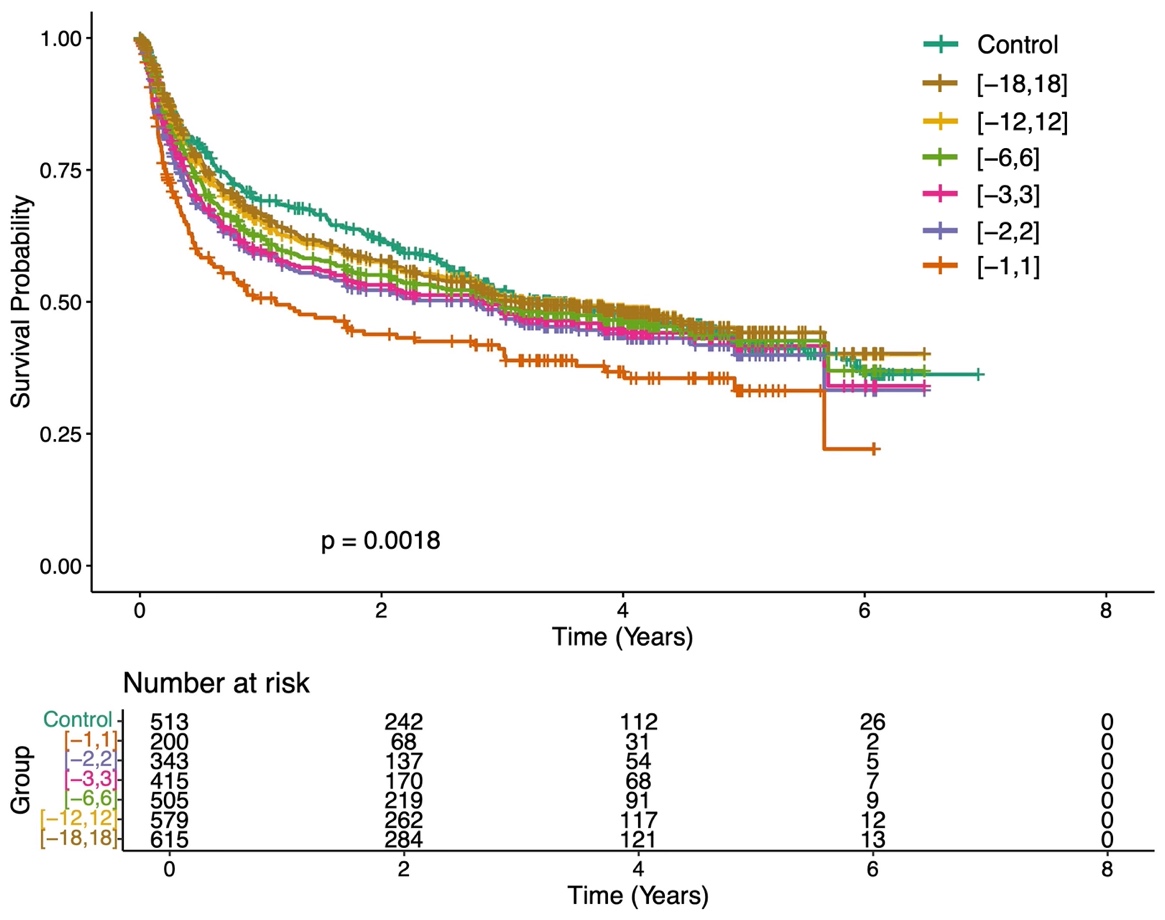

**B.** MGBD

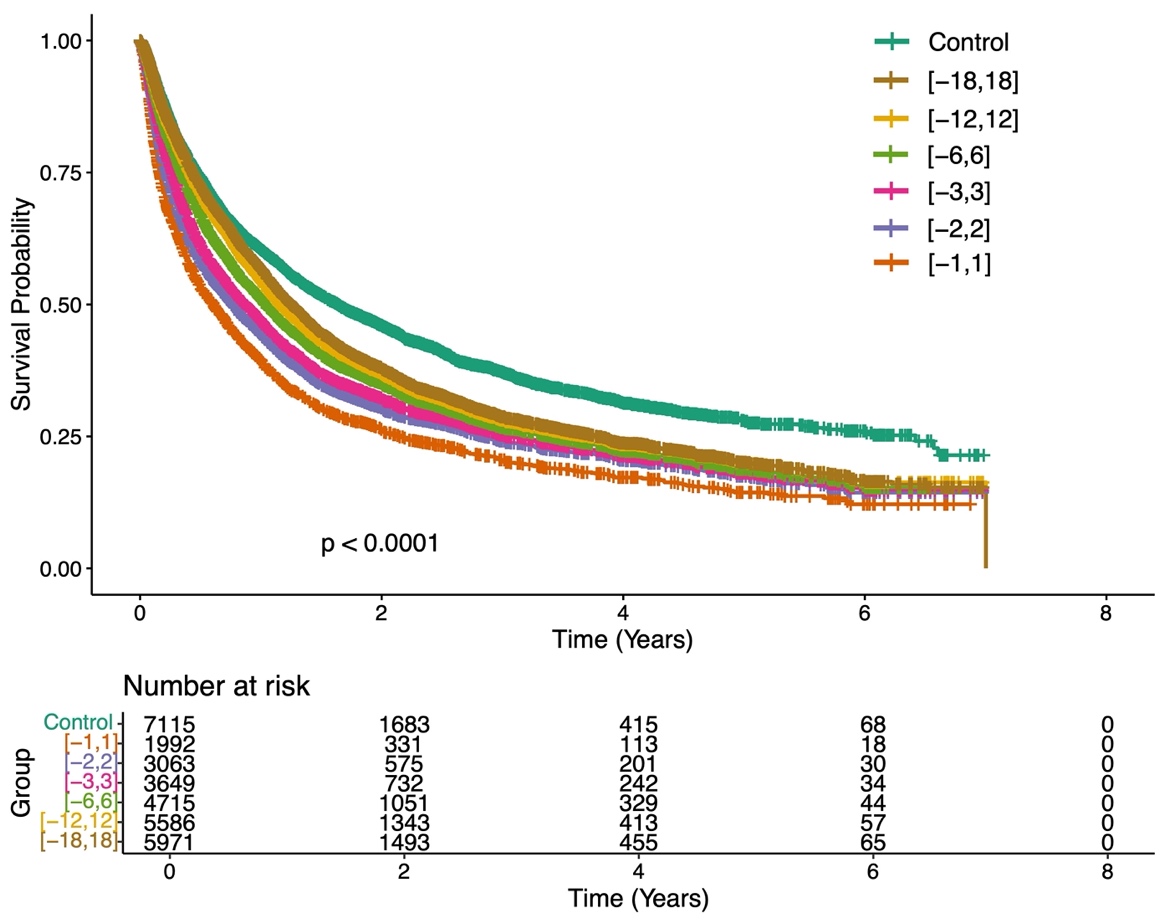

**C.** TriNetX

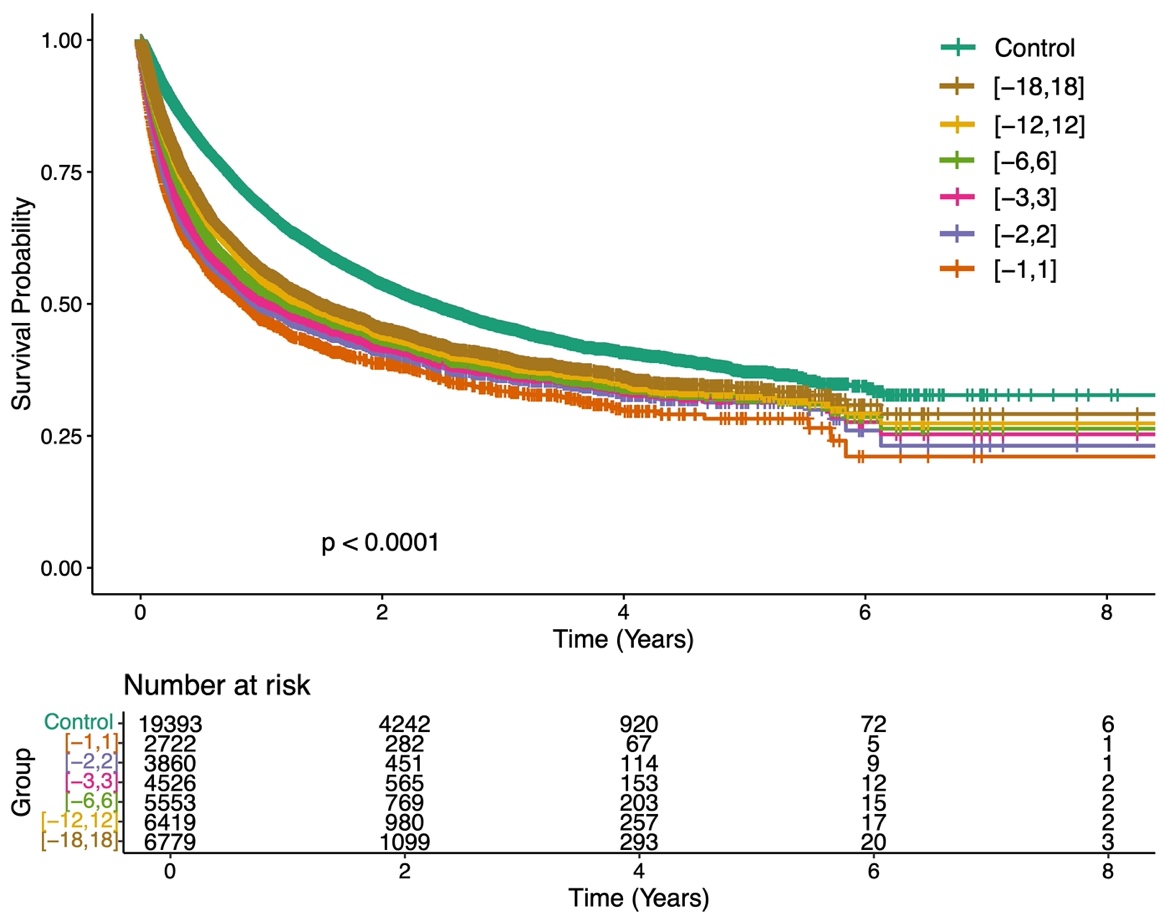

**Table S12.** Effect of systemic immunosuppression on overall survival using multivariable CoxPH modeling.

|  | | **Time Window from ICI Initiation (months)** | | | | | | |
| --- | --- | --- | --- | --- | --- | --- | --- | --- |
| **Cohort** |  | **[-12, 12]** | **[-6, 6]** | **[-5, 5]** | **[-4, 4]** | **[-3, 3]** | **[-2, 2]** | **[-1, 1]** |
| MGBD | HR  95% CI | 1.27  [1.17, 1.38] | 1.20  [1.12, 1.28] | 1.17  [1.09, 1.24] | 1.14  [1.07, 1.21] | 1.17  [1.10, 1.24] | 1.25  [1.18, 1.33] | 1.53  [1.43, 1.63] |
|  | P-value | < 0.0001 | < 0.0001 | < 0.0001 | < 0.0001 | < 0.0001 | < 0.0001 | < 0.0001 |
| TriNetX | HR  95% CI | 1.05  [0.95, 1.16] | 1.14  [1.01, 1.24] | 1.16  [1.07, 1.25] | 1.18  [1.10, 1.27] | 1.31  [1.22, 1.40] | 1.41  [1.32, 1.51] | 1.62  [1.51, 1.74] |
|  | P-value | 0.35 | 0.0008 | < 0.0001 | < 0.0001 | < 0.0001 | < 0.0001 | < 0.0001 |

Adjusted Hazard Ratios (HRs) were derived from multivariable Cox Ph models adjusted by sex, age at ICI initiation, race, ethnicity, cancer type, cancer stage, Charlson comorbidity index, ICI type, and ICI cycles.

CoxPH: Cox proportional hazards. HR: Hazard Ratio; CI: Confidence Interval.

**Table S13.** Effect of systemic immunosuppression on overall survival using multivariable AFT modeling.

|  | | **Time Window from ICI Initiation (months)** | | | | | | |
| --- | --- | --- | --- | --- | --- | --- | --- | --- |
| **Cohort** |  | **[-1, 1]** | **[-1, 2]** | **[-1, 3]** | **[-1, 4]** | **[-1, 5]** | **[-1, 6]** | **[-1, 12]** |
| MGBD | TR  95% CI | 0.49  [0.45, 0.54] | 0.63  [0.58, 0.69] | 0.72  [0.66, 0.79] | 0.71  [0.65, 0.78] | 0.70  [0.63, 0.76] | 0.69  [0.63, 0.76] | 0.67  [0.60, 0.75] |
|  | P-value | < 0.0001 | < 0.0001 | < 0.0001 | < 0.0001 | < 0.0001 | < 0.0001 | < 0.0001 |
| TriNetX | TR  95% CI | 0.44  [0.40, 0.49] | 0.50  [0.45, 0.55] | 0.55  [0.50, 0.61] | 0.62  [0.56, 1.48] | 0.66  [0.59, 0.73] | 0.65  [0.58, 0.73] | 0.79  [0.68, 0.92] |
|  | P-value | < 0.0001 | < 0.0001 | < 0.0001 | < 0.0001 | < 0.0001 | < 0.0001 | < 0.0001 |

Adjusted Time Ratios (TRs): TR < 1 means that the covariate (e.g., systemic glucocorticoid immunosuppression) is associated with shorter survival. Adjusted TRs were derived from multivariable log-normal AFT models and represented their exponentiated coefficients. The AFT models were adjusted by sex, age at ICI initiation, race, ethnicity, cancer type, cancer stage, Charlson comorbidity index, ICI type, and ICI cycles.

AFT: Accelerated Failure Time; TR: Time Ratio; CI: Confidence Interval; ICI: immune checkpoint inhibitor.

**Table S14.** The full AFT model for the association of sISP and overall survival with the [-1, 1] window (MGBD).

| **Characteristics** | **Time Ratio** | **95% CI** | **P-value** |
| --- | --- | --- | --- |
| **sISP** |  |  |  |
| No | Ref | Ref |  |
| Yes | 0.49 | 0.45, 0.54 | <0.0001 |
| **Sex** |  |  |  |
| Female | Ref | Ref |  |
| Male | 0.87 | 0.81, 0.94 | 0.00028 |
| **Race** |  |  |  |
| White | Ref | Ref |  |
| Black or African American | 0.92 | 0.75, 1.1 | 0.43 |
| Asian | 0.92 | 0.76, 1.1 | 0.39 |
| Other/Unavailable | 1.0 | 0.83, 1.2 | 0.9 |
| **Ethnicity** |  |  |  |
| Not Hispanic | Ref | Ref |  |
| Hispanic | 1.1 | 0.84, 1.3 | 0.6 |
| Unavailable | 0.83 | 0.72, 0.96 | 0.0097 |
| **Age at ICI Initiation (years)** |  |  |  |
| ≤60 | Ref | Ref |  |
| 61–70 | 0.90 | 0.82, 0.98 | 0.014 |
| 71–80 | 0.73 | 0.67, 0.80 | <0.0001 |
| >80 | 0.48 | 0.43, 0.55 | <0.0001 |
| **Charlson Comorbidity Index** |  |  |  |
| 0 | Ref | Ref |  |
| 1–2 | 1.0 | 0.63, 1.7 | 0.94 |
| 3–4 | 1.2 | 0.71, 1.9 | 0.55 |
| ≥5 | 0.84 | 0.52, 1.4 | 0.46 |
| **Cancer Type** |  |  |  |
| Thoracic | Ref | Ref |  |
| Male Genital/Urinary | 1.3 | 1.2, 1.5 | <0.0001 |
| Digestive | 0.60 | 0.53, 0.67 | <0.0001 |
| Melanoma | 3.5 | 3.0, 4.1 | <0.0001 |
| Other Skin Malignancy | 1.6 | 1.4, 1.9 | <0.0001 |
| Breast | 1.0 | 0.90, 1.2 | 0.54 |
| Lymphoid/Hematopoietic | 1.0 | 0.86, 1.2 | 0.87 |
| Female Genital | 0.91 | 0.77, 1.1 | 0.29 |
| Brain/Nervous/Eye | 0.57 | 0.48, 0.68 | <0.0001 |
| Oral/Lip/Pharynx | 0.87 | 0.72, 1.0 | 0.13 |
| Other | 0.95 | 0.83, 1.1 | 0.45 |
| **Cancer Stage** |  |  |  |
| Locoregional | Ref | Ref |  |
| Distant | 0.65 | 0.58, 0.72 | <0.0001 |
| **ICI Type** |  |  |  |
| CTLA-4 | Ref | Ref |  |
| PD-1 | 0.81 | 0.58, 1.1 | 0.23 |
| PD-L1 | 0.89 | 0.63, 1.3 | 0.51 |
| Combination | 0.75 | 0.52, 1.1 | 0.1 |
| **ICI Cycles** |  |  |  |
| 1 | Ref | Ref |  |
| 2 | 2.3 | 2.0, 2.5 | <0.0001 |
| 3 | 2.8 | 2.5, 3.2 | <0.0001 |
| **Non-ICI Treatment** |  |  |  |
| None | Ref | Ref |  |
| Conventional Chemotherapy | 0.61 | 0.57, 0.66 | <0.0001 |
| Targeted Therapy | 0.73 | 0.66, 0.82 | <0.0001 |

**Table S15.** The full AFT model for the association of sISP and overall survival with the [-3, 3] window (MGBD).

| **Characteristics** | **Time Ratio** | **95% CI** | **P-value** |
| --- | --- | --- | --- |
| **sISP** |  |  |  |
| No | Ref | Ref |  |
| Yes | 0.75 | 0.69, 0.83 | <0.0001 |
| **Sex** |  |  |  |
| Female | Ref | Ref |  |
| Male | 0.89 | 0.83, 0.97 | 0.0065 |
| **Race** |  |  |  |
| White | Ref | Ref |  |
| Black or African American | 0.90 | 0.73, 1.1 | 0.36 |
| Asian | 1.1 | 0.88, 1.3 | 0.45 |
| Other/Unavailable | 0.96 | 0.77, 1.2 | 0.68 |
| **Ethnicity** |  |  |  |
| Not Hispanic | Ref | Ref |  |
| Hispanic | 1.1 | 0.85, 1.4 | 0.49 |
| Unavailable | 0.86 | 0.74, 1.0 | 0.05 |
| **Age at ICI Initiation (years)** |  |  |  |
| ≤60 | Ref | Ref |  |
| 61–70 | 0.90 | 0.82, 0.99 | 0.024 |
| 71–80 | 0.77 | 0.70, 0.85 | <0.0001 |
| >80 | 0.46 | 0.40, 0.52 | <0.0001 |
| **Charlson Comorbidity Index** |  |  |  |
| 0 | Ref | Ref |  |
| 1–2 | 0.97 | 0.58, 1.6 | 0.91 |
| 3–4 | 1.1 | 0.66, 1.9 | 0.67 |
| ≥5 | 0.93 | 0.56, 1.6 | 0.79 |
| **Cancer Type** |  |  |  |
| Thoracic | Ref | Ref |  |
| Male Genital/Urinary | 1.4 | 1.2, 1.6 | <0.0001 |
| Digestive | 0.74 | 0.65, 0.85 | <0.0001 |
| Melanoma | 3.3 | 2.8, 3.8 | <0.0001 |
| Other Skin Malignancy | 1.5 | 1.3, 1.8 | <0.0001 |
| Breast | 1.1 | 0.95, 1.3 | 0.19 |
| Lymphoid/Hematopoietic | 1.2 | 0.98, 1.4 | 0.076 |
| Female Genital | 1.0 | 0.84, 1.2 | 0.97 |
| Brain/Nervous/Eye | 0.42 | 0.35, 0.51 | <0.0001 |
| Oral/Lip/Pharynx | 0.84 | 0.69, 1.0 | 0.085 |
| Other | 1.1 | 0.92, 1.2 | 0.41 |
| **Cancer Stage** |  |  |  |
| Locoregional | Ref | Ref |  |
| Distant | 0.59 | 0.53, 0.66 | <0.0001 |
| **ICI Type** |  |  |  |
| CTLA-4 | Ref | Ref |  |
| PD-1 | 0.87 | 0.62, 1.2 | 0.41 |
| PD-L1 | 0.85 | 0.60, 1.2 | 0.38 |
| Combination | 1.0 | 0.71, 1.4 | 0.96 |
| **ICI Cycles** |  |  |  |
| 1 | Ref | Ref |  |
| 2 | 0.98 | 0.82, 1.2 | 0.85 |
| 3 | 1.0 | 0.84, 1.2 | 0.92 |
| 4 | 1.8 | 1.5, 2.1 | <0.0001 |
| ≥5 | 3.1 | 2.6, 3.7 | <0.0001 |
| **Non-ICI Treatment** |  |  |  |
| None | Ref | Ref |  |
| Conventional Chemotherapy | 0.56 | 0.51, 0.61 | <0.0001 |
| Targeted Therapy | 0.66 | 0.59, 0.74 |  |

**Table S16.** The full AFT model for the association of sISP and overall survival with the [-1, 1] window (TriNetX).

| **Characteristics** | **Time Ratio** | **95% CI** | **P-value** |
| --- | --- | --- | --- |
| **sISP** |  |  |  |
| No | Ref | Ref |  |
| Yes | 0.44 | 0.40, 0.49 | <0.0001 |
| **Sex** |  |  |  |
| Female | Ref | Ref |  |
| Male | 0.89 | 0.83, 0.95 | 0.00052 |
| **Race** |  |  |  |
| White | Ref | Ref |  |
| Black or African American | 1.1 | 0.92, 1.3 | 0.27 |
| Asian | 1.4 | 1.1, 1.6 | 0.0013 |
| Other/Unavailable | 1.1 | 0.97, 1.3 | 0.12 |
| **Ethnicity** |  |  |  |
| Not Hispanic | Ref | Ref |  |
| Hispanic | 1.1 | 0.85, 1.4 | 0.49 |
| Unavailable | 0.86 | 0.74, 1.0 | 0.05 |
| **Age at ICI Initiation (years)** |  |  |  |
| ≤60 | Ref | Ref |  |
| 61–70 | 0.92 | 0.85, 0.99 | 0.026 |
| 71–80 | 0.84 | 0.77, 0.91 | <0.0001 |
| >80 | 0.52 | 0.47, 0.59 | <0.0001 |
| **Charlson Comorbidity Index** |  |  |  |
| 0 | Ref | Ref |  |
| 1–2 | 0.97 | 0.66, 1.4 | 0.89 |
| 3–4 | 0.73 | 0.49, 1.1 | 0.12 |
| ≥5 | 0.63 | 0.43, 0.92 | 0.017 |
| **Cancer Type** |  |  |  |
| Thoracic | Ref | Ref |  |
| Male Genital/Urinary | 1.3 | 1.2, 1.4 | <0.0001 |
| Digestive | 0.84 | 0.75, 0.93 | 0.00096 |
| Melanoma | 2.8 | 2.5, 3.1 | <0.0001 |
| Other Skin Malignancy | 1.7 | 1.5, 2.0 | <0.0001 |
| Breast | 1.1 | 0.96, 1.3 | 0.14 |
| Lymphoid/Hematopoietic | 1.1 | 0.95, 1.3 | 0.2 |
| Female Genital | 1.2 | 0.98, 1.4 | 0.077 |
| Brain/Nervous/Eye | 0.60 | 0.50, 0.72 | <0.0001 |
| Oral/Lip/Pharynx | 0.84 | 0.71, 0.99 | 0.036 |
| Other | 1.1 | 0.96, 1.2 | 0.2 |
| **Cancer Stage** |  |  |  |
| Locoregional | Ref | Ref |  |
| Distant | 0.61 | 0.56, 0.67 | <0.0001 |
| **ICI Type** |  |  |  |
| CTLA-4 | Ref | Ref |  |
| PD-1 | 1.1 | 0.85, 1.5 | 0.38 |
| PD-L1 | 1.7 | 1.3, 2.3 | 0.00053 |
| Combination | 1.2 | 0.87, 1.6 | 0.28 |
| **ICI Cycles** |  |  |  |
| 1 | Ref | Ref |  |
| 2 | 1.5 | 1.4, 1.6 | <0.0001 |
| 3 | 1.3 | 1.2, 1.5 | <0.0001 |
| **Non-ICI Treatment** |  |  |  |
| None | Ref | Ref |  |
| Conventional Chemotherapy | 0.55 | 0.51, 0.59 | <0.0001 |
| Targeted Therapy | 0.66 | 0.60, 0.73 | <0.0001 |

**Table S17.** The full AFT model for the association of sISP and overall survival with the [-3, 3] window (TriNetX).

| **Characteristics** | **Time Ratio** | **95% CI** | **P-value** |
| --- | --- | --- | --- |
| **sISP** | | |  |
| No | Ref | Ref |  |
| Yes | 0.62 | 0.56, 0.68 | <0.0001 |
| **Sex** |  |  |  |
| Female | Ref | Ref |  |
| Male | 0.90 | 0.84, 0.97 | 0.0037 |
| **Race** |  |  |  |
| White | Ref | Ref |  |
| Black or African American | 1.0 | 0.84, 1.3 | 0.77 |
| Asian | 1.4 | 1.2, 1.8 | 0.00035 |
| Other/Unavailable | 1.1 | 0.89, 1.3 | 0.5 |
| **Ethnicity** |  |  |  |
| Not Hispanic | Ref | Ref |  |
| Hispanic | 1.1 | 0.93, 1.4 | 0.2 |
| Unavailable | 1.2 | 1.1, 1.4 | 0.0064 |
| **Age at ICI Initiation (years)** |  |  |  |
| ≤60 | Ref | Ref |  |
| 61–70 | 0.88 | 0.81, 0.95 | 0.0019 |
| 71–80 | 0.79 | 0.72, 0.86 | <0.0001 |
| >80 | 0.54 | 0.47, 0.61 | <0.0001 |
| **Charlson Comorbidity Index** |  |  |  |
| 0 | Ref | Ref |  |
| 1–2 | 1.1 | 0.72, 1.6 | 0.68 |
| 3–4 | 0.85 | 0.56, 1.3 | 0.45 |
| ≥5 | 0.79 | 0.53, 1.2 | 0.26 |
| **Cancer Type** |  |  |  |
| Thoracic | Ref | Ref |  |
| Male Genital/Urinary | 1.3 | 1.1, 1.4 | <0.0001 |
| Digestive | 0.94 | 0.83, 1.1 | 0.29 |
| Melanoma | 2.7 | 2.4, 3.1 | <0.0001 |
| Other Skin Malignancy | 1.6 | 1.4, 1.9 | <0.0001 |
| Breast | 1.2 | 1.0, 1.4 | 0.03 |
| Lymphoid/Hematopoietic | 1.3 | 1.1, 1.5 | 0.0019 |
| Female Genital | 1.1 | 0.93, 1.3 | 0.26 |
| Brain/Nervous/Eye | 0.58 | 0.48, 0.70 | <0.0001 |
| Oral/Lip/Pharynx | 0.80 | 0.67, 0.96 | 0.016 |
| Other | 1.1 | 0.95, 1.2 | 0.23 |
| **Cancer Stage** |  |  |  |
| Locoregional | Ref | Ref |  |
| Distant | 0.54 | 0.49, 0.60 | <0.0001 |
| **ICI Type** |  |  |  |
| CTLA4 | Ref | Ref |  |
| PD1 | 1.0 | 0.75, 1.4 | 0.94 |
| PDL1 | 1.3 | 0.97, 1.8 | 0.073 |
| Combination | 1.4 | 1.0, 1.9 | 0.04 |
| **ICI Cycles** |  |  |  |
| 1 | Ref | Ref |  |
| 2 | 0.84 | 0.73, 0.96 | 0.012 |
| 3 | 0.82 | 0.72, 0.93 | 0.0022 |
| 4 | 1.1 | 1.0, 1.3 | 0.022 |
| ≥5 | 1.8 | 1.6, 2.0 | <0.0001 |
| **Non-ICI Treatment** |  |  |  |
| None | Ref | Ref |  |
| Conventional Chemotherapy | 0.53 | 0.49, 0.57 | <0.0001 |
| Targeted Therapy | 0.64 | 0.57, 0.71 | <0.0001 |

**Table S18.** Multivariable AFT modeling of systemic immunosuppression on overall survival (Manual cohort)

|  | | **Time Window from ICI Initiation (months)** | | | | | |
| --- | --- | --- | --- | --- | --- | --- | --- |
| **Matching** |  | **[-6, 6]** | **[-5, 5]** | **[-4, 4]** | **[-3, 3]** | **[-2, 2]** | **[-1, 1]** |
| No | TR  95% CI | 0.92  [0.62, 1.4] | 0.78  [0.55, 1.1] | 0.75  [0.52,1.1] | 0.64  [0.45, 0.92] | 0.65  [0.45, 0.93] | 0.32  [0.22,0.48] |
|  | P-value | 0.69 | 0.18 | 0.14 | 0.017 | 0.018 | <0.0001 |
| 1:2 | TR  95% CI | 0.92  [0.62, 1.4] | 0.78  [0.55, 1.1] | 0.75  [0.52, 1.1] | 0.64  [0.45, 0.92] | 0.66  [0.45, 0.95] | 0.31  [0.20, 0.49] |
|  | P-value | 0.7 | 0.2 | 0.14 | 0.0017 | 0.026 | <0.0001 |

TR: Time Ratio; 95% CI: 95% Confidence Interval.

**Table S19.** Multivariable AFT modeling of systemic immunosuppression on overall survival (1:2 matching)

|  | | **Time Window from ICI Initiation (months)** | | | | | | |
| --- | --- | --- | --- | --- | --- | --- | --- | --- |
| **Cohort** |  | **[-12, 12]** | **[-6, 6]** | **[-5, 5]** | **[-4, 4]** | **[-3, 3]** | **[-2, 2]** | **[-1, 1]** |
| MGBD | TR  95% CI | 0.71  [0.64, 0.79] | 0.75  [0.69, 0.82] | 0.77  [0.70, 0.84] | 0.79  [0.72, 0.86] | 0.77  [0.71, 0.84] | 0.70  [0.64, 0.77] | 0.50  [0.45, 0.56] |
|  | P-value | <0.0001 | <0.0001 | <0.0001 | <0.0001 | <0.0001 | <0.0001 | <0.0001 |
| TriNetX | TR  95% CI | 0.83  [0.71,0.98] | 0.75  [0.66,0.86] | 0.72  [0.64,0.81] | 0.71  [0.63, 0.79] | 0.59  [0.52, 0.66] | 0.55  [0.49, 0.62] | 0.47  [0.40, 0.54] |
|  | P-value | 0.041 | <0.0001 | <0.0001 | <0.0001 | <0.0001 | <0.0001 | <0.0001 |

TR: Time Ratio; CI: Confidence Interval.

**Figure S3.** Landmark survival analysis for the association of systemic immunosuppression with overall survival (Manual cohort).

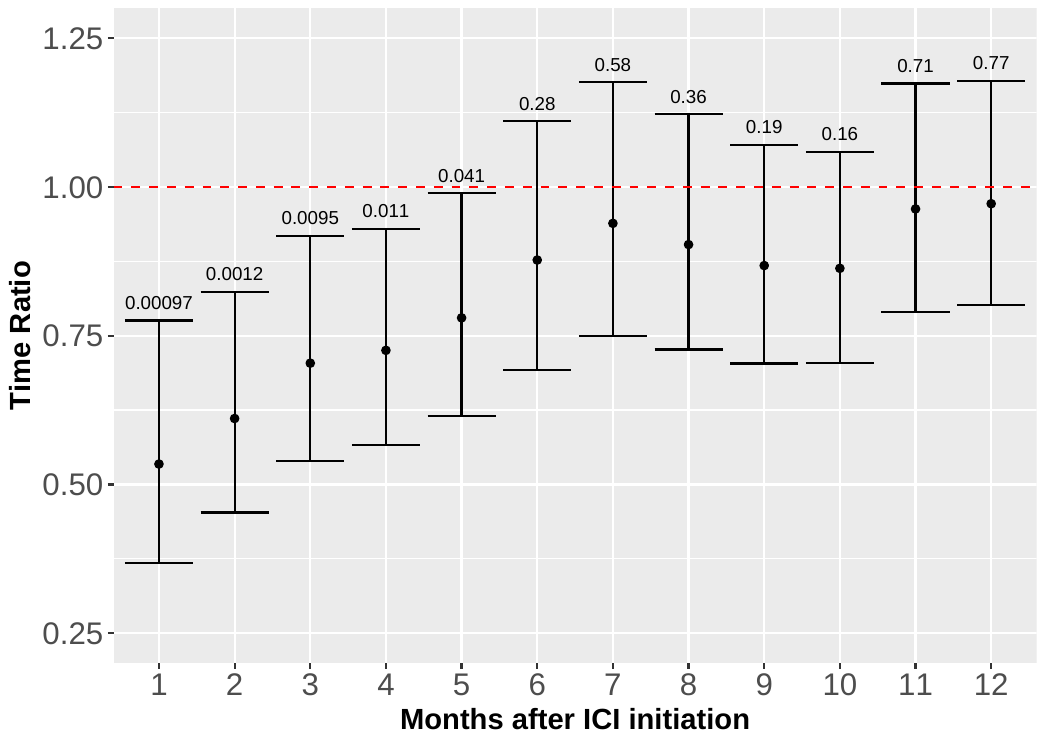

**Table S20.** Indications for systemic immunosuppression (Manual).

| **Window (months)**  **Patients with sISP (N)** | **[-6, 6]**  **505** | **[-5, 5]**  **483** | **[-4, 4]**  **456** | **[-3, 3]**  **415** | **[-2, 2]**  **343** | **[-1, 1]**  **200** |
| --- | --- | --- | --- | --- | --- | --- |
| **irAEs** | 171  (33.9%) | 157  (32.5%) | 146  (32%) | 131  (31.6%) | 93  (27.1%) | 43  (21.5%) |
| **Cancer palliation** | 131  (25.9%) | 127  (26.3%) | 123  (27.0%) | 112  (27.0%) | 97  (28.3%) | 66  (33.0%) |
| **Pre-existing condition** | 29  (5.7%) | 30  (6.2%) | 29  (6.4%) | 21  (5.1%) | 20  (5.8%) | 19  (9.5%) |
| **Other** | 174  (34.5%) | 169  (35%) | 158  (34.7%) | 151  (36.4%) | 133  (38.8%) | 72  (36%) |

**irAEs:** includes patients who received systemic immunosuppression for managing irAEs and/or other conditions.

**Cancer palliation:** includes patients who received systemic immunosuppression for cancer palliation and/or other conditions, except irAEs.

**Pre-existing condition:** includes patients who received systemic immunosuppression for managing pre-existing and/or other conditions, except irAEs and cancer palliation.

**Other:** includes patients who received systemic immunosuppression for conditions not mentioned above, such as transplantation.

**Figure S4.** Reasons for immunosuppression with different dosage and duration categories (Manual cohort).

**A.**

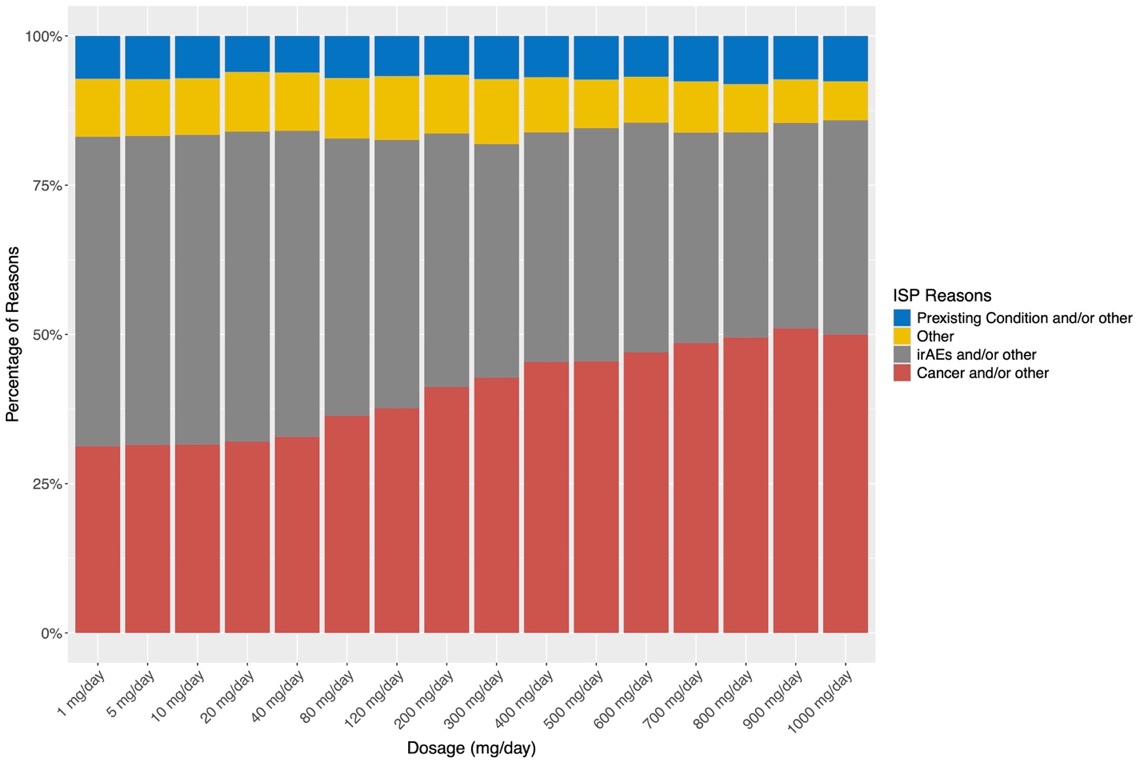

B.

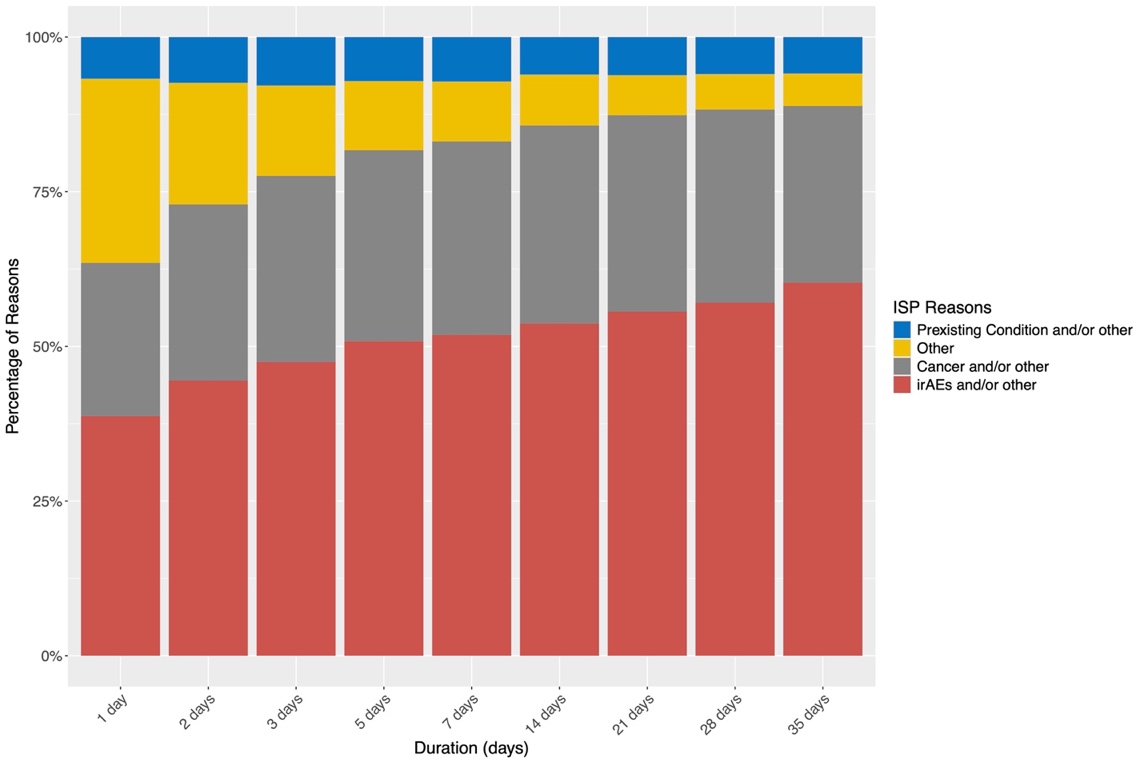

**A**. Breakdown of immunosuppression indications across dosage categories. Indications are categorized into 4 types: “Preexisting Condition and/or Other”, “Cancer and/or Other”, “irAEs and/or Other”, and “Other”. Dosage is presented in prednisone equivalent (Table S7).

**B**. Breakdown of immunosuppression indications across duration categories.

**Figure S5.** Reasons for immunosuppression across different dosage and duration categories in patients exclusively treated with glucocorticoids (Manual Cohort)

A.

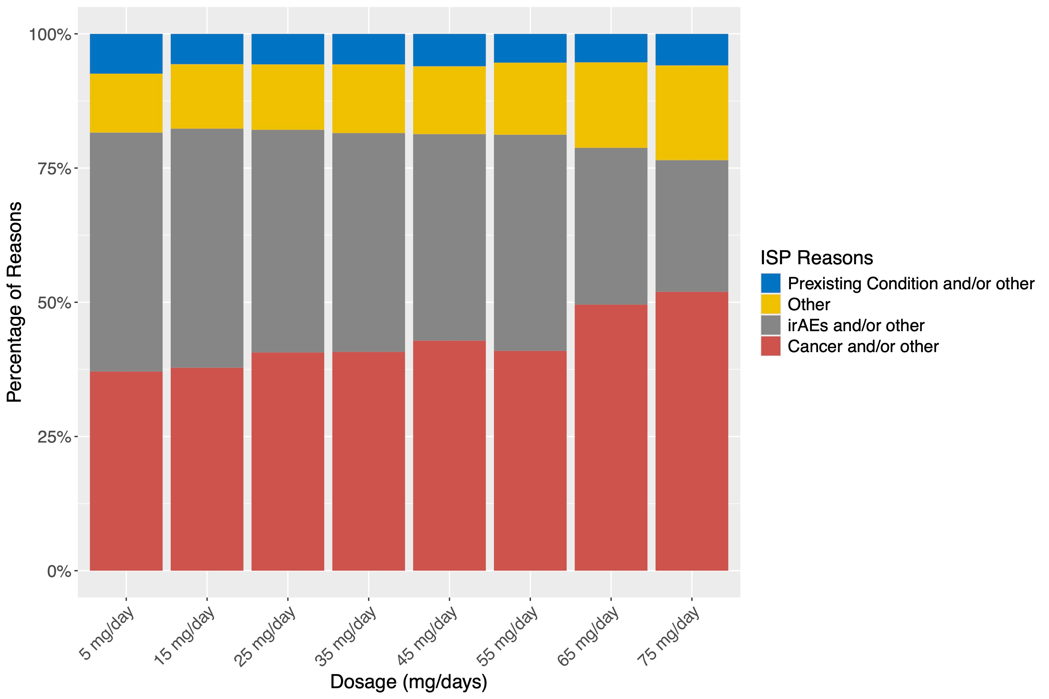

B.

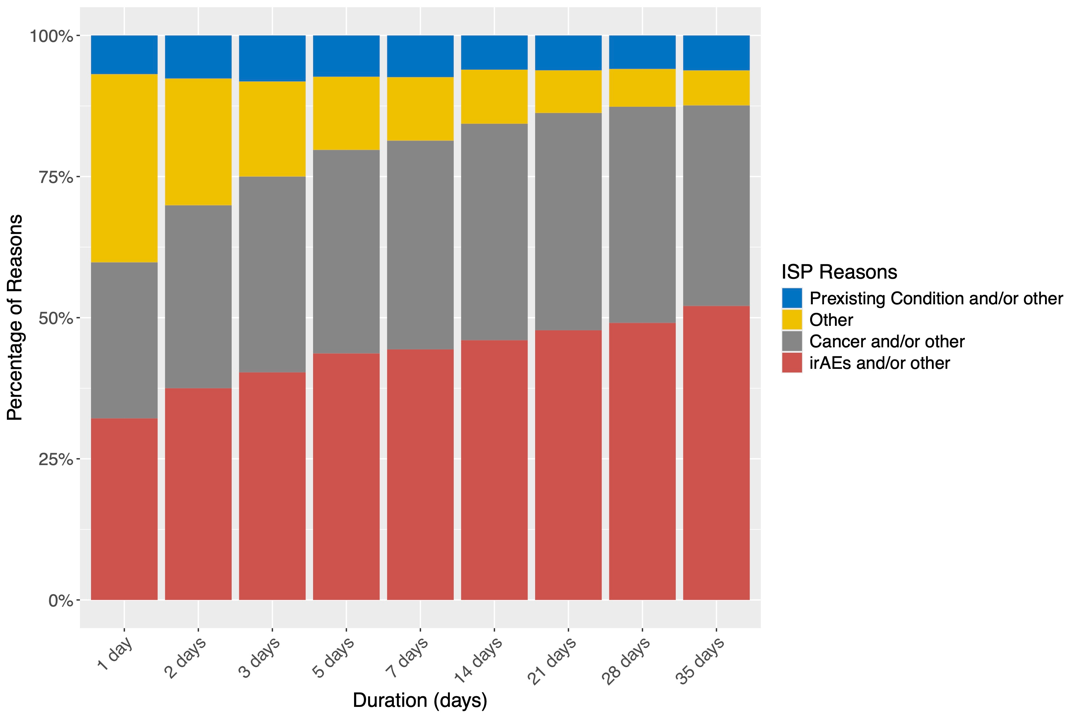

A. Breakdown of immunosuppression indications across dosage categories. Indications are categorized into 4 types: “Preexisting Condition and/or Other”, “Cancer and/or Other”, “irAEs and/or Other”, and “Other”. Dosage is presented in prednisone equivalent.

B. Breakdown of ISP indications across duration categories.

**Table S21.** Characteristics of irAE patients stratified by sISP status (MGBD).

| **Characteristics** | **sISP**  (N=1,038) | **No sISP**  (N=2,246) | **P-value** |
| --- | --- | --- | --- |
| **Sex** |  |  |  |
| Female | 484 (46.6%) | 951 (42.3%) | 0.024 |
| Male | 554 (53.4%) | 1,295 (57.7%) |  |
| **Race** |  |  |  |
| White | 953 (91.8%) | 2,059 (91.7%) | 0.83 |
| Black or African American | 28 (2.7%) | 54 (2.4%) |  |
| Asian | 22 (2.1%) | 58 (2.6%) |  |
| Other/Unavailable | 35 (3.4%) | 75 (3.3%) |  |
| **Ethnicity** |  |  |  |
| Not Hispanic | 964 (92.9%) | 2,040 (90.8%) | 0.15 |
| Hispanic | 24 (2.3%) | 64 (2.8%) |  |
| Unavailable | 50 (4.8%) | 142 (6.3%) |  |
| **Age at ICI Initiation (years)** |  |  |  |
| ≤60 | 321 (30.9%) | 577 (25.7%) | 0.0026 |
| 61–70 | 309 (29.8%) | 681 (30.3%) |  |
| 71–80 | 282 (27.2%) | 631 (28.1%) |  |
| >80 | 126 (12.1%) | 357 (15.9%) |  |
| **Charlson Comorbidity Index** |  |  |  |
| 0 | 0 (0%) | 21 (0.9%) | 0.00031 |
| 1–2 | 83 (8.0%) | 254 (11.3%) |  |
| 3–4 | 67 (6.5%) | 142 (6.3%) |  |
| ≥5 | 888 (85.5%) | 1,829 (81.4%) |  |
| **Cancer Type** |  |  |  |
| Thoracic | 231 (22.3%) | 487 (21.7%) | 0.0049 |
| Male Genital/Urinary | 120 (11.6%) | 304 (13.5%) |  |
| Digestive | 100 (9.6%) | 251 (11.2%) |  |
| Melanoma | 229 (22.1%) | 438 (19.5%) |  |
| Other Skin Malignancy | 69 (6.6%) | 179 (8.0%) |  |
| Breast | 30 (2.9%) | 68 (3.0%) |  |
| Lymphoid/Hematopoietic | 52 (5.0%) | 85 (3.8%) |  |
| Female Genital | 26 (2.5%) | 82 (3.7%) |  |
| Brain/Nervous/Eye | 52 (5.0%) | 64 (2.8%) |  |
| Oral/Lip/Pharynx | 61 (5.9%) | 112 (5.0%) |  |
| Other | 68 (6.6%) | 176 (7.8%) |  |
| **Cancer Stage** |  |  |  |
| Locoregional | 145 (14.0%) | 485 (21.6%) | < 0.0001 |
| Distant | 893 (86.0%) | 1,761 (78.4%) |  |
| **Non-ICI Treatment** |  |  |  |
| Conventional Chemotherapy | 142 (13.7%) | 338 (15.0%) | 0.49 |
| Targeted Therapy | 35 (3.4%) | 84 (3.7%) |  |
| None | 861 (82.9%) | 1,824 (81.2%) |  |
| **ICI Type** |  |  |  |
| PD-1 | 720 (69.4%) | 1,752 (78.0%) | < 0.0001 |
| PD-L1 | 67 (6.5%) | 233 (10.4%) |  |
| CTLA-4 | 33 (3.2%) | 23 (1.0%) |  |
| Combination | 218 (21.0%) | 238 (10.6%) |  |
| **ICI Treatment Cycles** |  |  |  |
| 1 | 155 (14.9%) | 129 (5.7%) | < 0.0001 |
| 2 | 208 (20.0%) | 164 (7.3%) |  |
| 3 | 178 (17.1%) | 231 (10.3%) |  |
| 4 | 213 (20.5%) | 459 (20.4%) |  |
| ≥5 | 284 (27.4%) | 1,263 (56.2%) |  |
| **Mortality status** |  |  |  |
| Alive | 462 (44.5%) | 1,350 (60.1%) | < 0.0001 |
| Dead | 576 (55.5%) | 896 (39.9%) |  |
| **Duration of Follow-up (days)** |  |  |  |
| Median [IQR] | 359 [113, 979] | 613 [272, 1100] | < 0.0001 |

**Table S22.** Characteristics of irAE patients stratified by sISP status (TriNetX).

| **Characteristics** | **sISP**  (N=1,299) | **No sISP**  (N=4,239) | **P-value** |
| --- | --- | --- | --- |
| **Sex** |  |  |  |
| Female | 537 (41.3%) | 1,782 (42.0%) | 0.68 |
| Male | 762 (58.7%) | 2,457 (58.0%) |  |
| **Race** |  |  |  |
| White | 1,216 (93.6%) | 3,834 (90.4%) | 0.0052 |
| Black or African American | 22 (1.7%) | 122 (2.9%) |  |
| Asian | 26 (2.0%) | 122 (2.9%) |  |
| Other/Unavailable | 35 (2.7%) | 161 (3.8%) |  |
| **Ethnicity** |  |  |  |
| Not Hispanic | 1,234 (95.0%) | 3,844 (90.7%) | < 0.0001 |
| Hispanic | 30 (2.3%) | 147 (3.5%) |  |
| Unavailable | 35 (2.7%) | 248 (5.9%) |  |
| **Age at ICI Initiation (years)** |  |  |  |
| ≤60 | 450 (34.6%) | 1,223 (28.9%) | 0.00012 |
| 61–70 | 400 (30.8%) | 1,290 (30.4%) |  |
| 71–80 | 326 (25.1%) | 1,246 (29.4%) |  |
| >80 | 123 (9.5%) | 480 (11.3%) |  |
| **Charlson Comorbidity Index** |  |  |  |
| 0 | 9 (0.7%) | 25 (0.6%) | 0.00044 |
| 1–2 | 100 (7.7%) | 482 (11.4%) |  |
| 3–4 | 81 (6.2%) | 318 (7.5%) |  |
| ≥5 | 1,109 (85.4%) | 3,414 (80.5%) |  |
| **Cancer Type** |  |  |  |
| Thoracic | 345 (26.6%) | 894 (21.1%) | < 0.0001 |
| Male Genital/Urinary | 213 (16.4%) | 733 (17.3%) |  |
| Digestive | 95 (7.3%) | 432 (10.2%) |  |
| Melanoma | 295 (22.7%) | 849 (20.0%) |  |
| Other Skin Malignancy | 74 (5.7%) | 292 (6.9%) |  |
| Breast | 27 (2.1%) | 86 (2.0%) |  |
| Lymphoid/Hematopoietic | 57 (4.4%) | 165 (3.9%) |  |
| Female Genital | 30 (2.3%) | 174 (4.1%) |  |
| Brain/Nervous/Eye | 35 (2.7%) | 92 (2.2%) |  |
| Oral/Lip/Pharynx | 35 (2.7%) | 197 (4.6%) |  |
| Other | 93 (7.2%) | 325 (7.7%) |  |
| **Cancer Stage** |  |  |  |
| Locoregional | 184 (14.2%) | 846 (20.0%) | < 0.0001 |
| Distant | 1,115 (85.8%) | 3,393 (80.0%) |  |
| **Non-ICI Treatment** |  |  |  |
| Conventional Chemotherapy | 201 (15.5%) | 588 (13.9%) | 0.31 |
| Targeted Therapy | 57 (4.4%) | 176 (4.2%) |  |
| None | 1,041 (80.1%) | 3,475 (82.0%) |  |
| **ICI Type** |  |  |  |
| PD-1 | 863 (66.4%) | 3,349 (79.0%) | < 0.0001 |
| PD-L1 | 102 (7.9%) | 381 (9.0%) |  |
| CTLA-4 | 37 (2.8%) | 48 (1.1%) |  |
| Combination | 297 (22.9%) | 461 (10.9%) |  |
| **ICI Treatment Cycles** |  |  |  |
| 1 | 236 (18.2%) | 465 (11.0%) | < 0.0001 |
| 2 | 204 (15.7%) | 339 (8.0%) |  |
| 3 | 233 (17.9%) | 493 (11.6%) |  |
| 4 | 258 (19.9%) | 949 (22.4%) |  |
| ≥5 | 368 (28.3%) | 1,993 (47.0%) |  |
| **Mortality status** |  |  |  |
| Alive | 808 (62.2%) | 3,021 (71.3%) | < 0.0001 |
| Dead | 491 (37.8%) | 1,218 (28.7%) |  |
| **Duration of Follow-up (days)** |  |  |  |
| Median [IQR] | 340 [119, 816] | 532 [242, 1050] | < 0.0001 |

**Figure S6.** Landmark survival analyses for the association between systemic immunosuppression and overall survival among irAE patients.

A. MGBD – Patients with irAEs

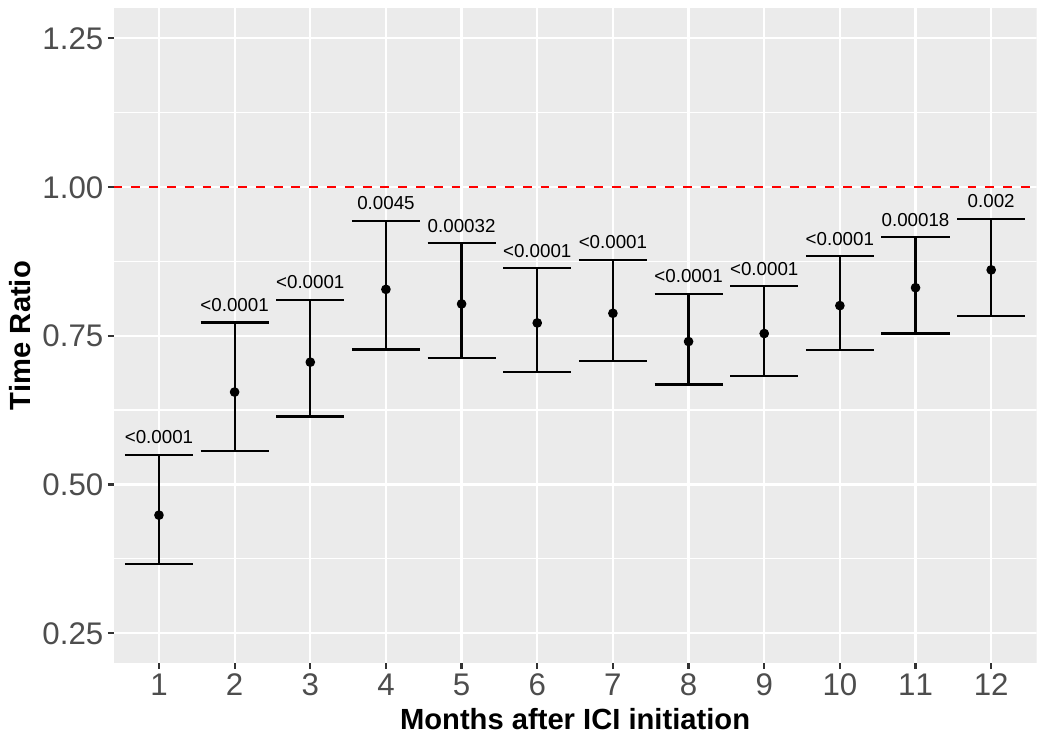

B. TriNetX – Patients with irAEs

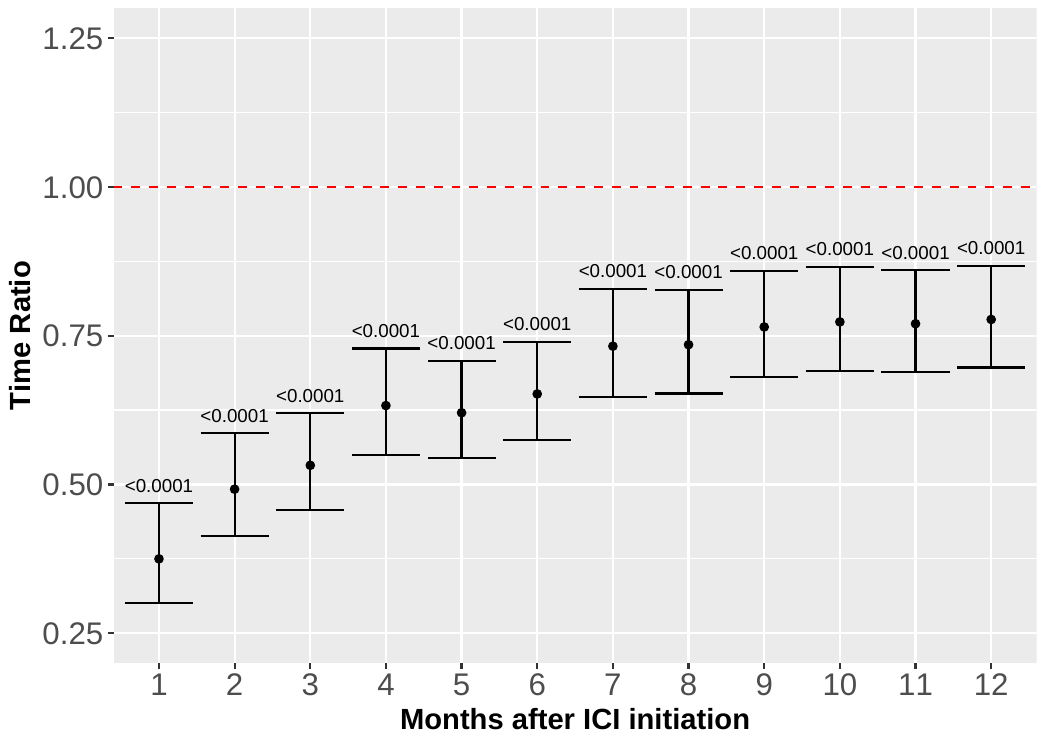
